# Circulating Metabolites Associated with Prothrombin Time and Activated Clotting Time in Children with Congenital Heart Disease: An Untargeted Metabolomics Study

**DOI:** 10.1101/2025.11.07.25339794

**Authors:** Shengxu Li, Dave Watson, Alissa Jorgenson, Zainab Adelekan, Kathleen Garland, Benjamin Deonovic, Leah Zupancich, Weihong Tang, David M. Overman, Marnie T. Huntley

**Author notes:** Correspondence (S.L.) and (M.T.H.).

## Abstract

**Background:** Coagulation markers are used to guide clinical anticoagulation decisions. We aimed to identify circulating metabolites that are associated with coagulation markers in children with congenital heart disease (CHD).

**Methods:** Plasma samples were separated from whole blood under consistent conditions. Untargeted metabolomic data were measured in plasma from up to 203 young patients (age range: 0 days-24 years) with CHD before cardiac surgery. Coagulation markers included activated partial thromboplastin time (aPTT), prothrombin time (PT), and activated clotting time (ACT). Weighted Gene Co-expression Network Analysis (WGCNA) was performed to explore metabolite modules (clusters). Associations of metabolites with the coagulation markers were assessed cross-sectionally with regression models, with false discovery rate (FDR) correction for multiple comparison. Associations between coagulation markers and ‘eigenmetabolites’ from WGCNA modules were assessed by correlation analysis.

**Results:** A total of 776 metabolites were included in the final analysis. Among these, 20 metabolites were associated with PT and one (valine) with ACT (FDR q value < 0.05). Among the metabolites associated with PT, the top three were retinol, 1-palmitoyl-GPI (16:0), and X-25371 (identity unknown). One module from WGCNA with metabolites from the lipid super pathway was correlated with PT (P = 0.004).

**Conclusion:** In this first attempt to identify novel metabolites for coagulation markers, we report 21 metabolites associated with PT or ACT in children with CHD. Future studies are needed to replicate these findings in independent cohorts and to elucidate the biological mechanisms linking these metabolites to hemostatic regulation.

## Introduction

Thrombosis and bleeding are common after heart surgery in children with congenital heart disease (CHD)[1,2]. Both thrombosis and bleeding increase risk of death and prolong the length of hospital stay in these children. Coagulation markers are important clinical indicators that guide clinical anticoagulation regimens. However, the hemostatic system in young children is still developing, and coagulation markers are known to be less correlated with hemostasis in children than in adults[3,4]. This exacerbates the challenges of designing effective anticoagulation strategies in children with CHD who are scheduled to have corrective heart surgery. Furthermore, evidence-based clinical guidelines for preventing and treating thrombosis and bleeding in children with CHD are lacking[2,5].

Despite these challenges, coagulation markers in children with CHD are still widely used for anticoagulation decision-making. Improving anticoagulation management in children with CHD requires a better understanding of the biological factors that influence commonly used coagulation markers. However, factors that may influence the measurement of these individual markers in children are not well understood. In recent years, metabolomic technology, which simultaneously measures up to tens of thousands of metabolites in biofluids, has made it possible to detect metabolic alterations and to elucidate new metabolic mechanisms on a hypothesis-free basis[6]. The non-targeted metabolomic approach—mass spectrometry coupled with liquid chromatography or gas chromatography—has been increasingly used to study complex pathophysiological processes[6]. In our previous report, we investigated metabolites associated with postoperative thrombosis, a clinically important outcome occurring after surgery[7]. Using the same cohort, the current study focused on cross-sectional associations between circulating metabolites and routinely measured coagulation markers before surgery.

## Materials and Methods

### Study sample

Young patients (aged 0-24 years) with CHD who planned to have corrective cardiac surgery at the Cardiovascular Care Center at Children’s Minnesota were eligible for the study. Patients with the following conditions were excluded: 1) body weight < 1800 grams at the time of surgery; 2) receiving extracorporeal membrane oxygenation at the time of surgery; 3) known clotting or bleeding disorder (e.g., Von Willebrand disease, Factor V Leiden, Factor VIII or Factor IX deficiency, methylenetetrahydrofolate reductase gene mutation); 4) isolated patent ductus arteriosus ligation; 5) parent or legal guardian unable to provide informed consent due to diminished capacity; and 6) having a second surgery during the same hospitalization.

Data and blood sample collection began in December 2019 and ended in August 2022.

### Clinical data collection

Clinical data, including demographics, diagnoses, pre-operation treatments, medical history (i.e., thrombosis or bleeding events), coagulation markers, and other relevant laboratory tests, were retrieved from the electronic medical record by A.J. and Z.A. The coagulation markers in the current study included activated partial thromboplastin time (aPTT), prothrombin time (PT), and activated clotting time (ACT). These markers were measured as part of routine clinical care. Samples for aPTT and PT were collected up to a month before surgery, but only those drawn within one week of surgery were included for analysis to ensure the samples were approximately contemporaneous. In contrast, ACT was sampled on the day of or one day prior to surgery.

### Blood sample collection and processing

Blood samples were collected right before incision across the study period. Immediately after the induction of anesthesia and before incision, 0.5-1.0 ml of whole blood was drawn through a central line into an EDTA tube, which was then inverted 8-10 times to assure anticoagulation. The blood sample was immediately put into an ice bag and transferred to the lab at Children’s Minnesota for immediate separation of plasma. A laboratory technician further inverted the tube manually a minimum of 20 times prior to centrifuging the sample for 10-15 minutes at a minimum of 2000 g. Separated plasma was aliquoted into a pre-chilled, barcoded cryovial (0.7 mL) provided by Metabolon (Morrisville, NC, a service provider for the study). Plasma samples were flash-frozen at -80°C before being shipped to Metabolon for metabolomic quantification.

### Metabolomic quantification

Metabolon quantified all plasma samples, which were shipped in two separate batches. Metabolon used the untargeted, ultrahigh performance liquid chromatography-tandem mass spectroscopy-based metabolomic quantification protocol and followed rigorous protocols for sample handling, processing, and metabolites profiling, including the use of internal standards, blank samples, and technical replicates. Metabolon applied several procedures to the raw metabolite data: first, the data were batch-adjusted by scaling raw metabolite levels to have a within-batch median of 1, then missing data were imputed to the median, and finally a log transformation was applied. In addition to chemical names of quantified metabolites, Metabolon-specific chemical ID, along with the Human Metabolome Database (HMDB) (https://www.hmdb.ca/) ID, was used to facilitate data analyses. Some metabolites did not have an HMDB ID.

We performed additional quality assurance via 5 within-batch and 10 between-batch duplicate samples, which were blinded to Metabolon. Spearman correlations were calculated between all duplicate groups for each metabolite. Only metabolites with call rates (i.e., non-missing data rates) ≥ 75% and with all correlation coefficients ≥ 0.5 were included for analysis. Among duplicate pairs, one sample was randomly selected for analysis.

## Statistical analysis

### Individual metabolite analysis

Multiple linear regression models were used to examine the associations of individual metabolites with each coagulation marker, while adjusting for age, race, sex, personal history of thrombosis, personal history of postoperative bleeding, oxygen saturation, hemoglobin, anticoagulation medication, and STAT (Society of Thoracic Surgeons-European Association for Cardio-Thoracic Surgery) score, which ranks the complexity of the planned surgery. For age adjustment, we used piecewise segmented regression with hinge points at 30 days, 1 year, and 10 years of age, which allowed us to capture non-linear associations across the entire time span of childhood while also capturing linear trends among age groups. For coagulation markers, PT, aPTT, and ACT were natural log-transformed to increase normality. We normalized the batch-adjusted, median-imputed, and natural log-transformed concentration of individual metabolites to z-scores before further analysis, so regression effect sizes are interpretable as the predicted change per one standard deviation increase on the log scale of concentration. Robust standard errors were used to mitigate potential impact of heteroscedasticity and non-normally distributed residual errors. To control for multiple comparisons, the positive false discovery rate (FDR) was controlled at the 5% level for each marker separately, specifically using a 0.05 significance threshold for the q-value—the FDR analogue of the p-value[8,9]. The FDR was controlled within each coagulation outcome, meaning that among the metabolites identified as significantly associated with a coagulation marker (e.g., PT), we expect at most 5% to be false positives. Regression models and Q-values were computed using SAS Enterprise Guide software (version 7.12, Cary, NC, USA), specifically the ‘GENMOD’ procedure and the ‘MULTTEST’ procedure with the ‘PFDR’ option, respectively.

### Weighted Gene Co-expression Network Analysis (WGCNA)

We performed WGCNA to identify modules (or clusters) of metabolites that were correlated together[10]. Metabolomic data are well suited for such analysis because metabolites in the body are inherently and closely correlated across interconnected metabolic pathways. WGCNA was used to detect and define modules of metabolites based on pair-wise correlations. To account for confounding factors, concentrations of individual metabolites were regressed on the same set of covariates used for adjustment in the single metabolite analysis (i.e., age, race, sex, etc.), and residuals from these regressions were used for pairwise correlations. Each module was summarized at a patient level using ‘eigenmetabolites,’ the first principal component of the metabolites within a given module, and these eigenmetabolites were then examined for correlation with coagulation markers. This analysis was performed via the ‘WGCNA’ library in R (version 4.3.0)[10-12].

## Results

From December 2019 to August 2022, 203 patients aged 0-24 years were enrolled with unique surgical encounters. Characteristics and coagulation markers of the study sample are shown in **Table 1**. Among this cohort, 157 and 158 patients had aPTT and PT values, respectively, sampled within one week of surgery; ACT data was available for 183 patients. A detailed quality control procedure for metabolomic data was described previously[7]. Our final dataset included 776 metabolites that met our filtering criteria.

**Table 1.** Characteristics of the study sample.

| Variable | n = 203 |
| --- | --- |
| Age years, median (Q1, Q3) | 1.4 (0.4-6.9) |
| Age group, n (%) |  |
| < 30 days | 20 (9.8) |
| 31-365 days | 78 (38.4) |
| 1-9 years | 62 (30.5) |
| > 10 years | 43 (21.2) |
| Female, n (%) | 87 (42.9) |
| Race, n (%) |  |
| White | 154 (75.9) |
| Black | 15 (7.4) |
| Other | 34 (16.7) |
| Medical history of bleeding, n (%) | 9 (4.4) |
| Medical history of thrombosis, n (%) | 31 (15.3) |
| STAT score, n (%) |  |
| 1 | 70 (34.5) |
| 2-3 | 94 (46.3) |
| 4-5 | 39 (19.2) |
| Pre-operative anticoagulant medication, n (%) | 55 (27.1) |
| Pre-operative O<sub>2</sub> saturation ≥ 95%, n (%) | 123 (60.6) |
| Hemoglobin (mg/dL), median (Q1, Q3) | 13.2 (12.0, 14.7) |
| aPTT seconds, median (Q1, Q3) (n=157) | 26.4 (24.9, 28.7) |
| PT seconds, median (Q1, Q3) (n=158) | 11.3 (10.8, 12.1) |
| ACT seconds, median (Q1, Q3) (n=183) | 158 (145, 169) |

ACT: activated clotting time; aPTT: activated partial thromboplastin time; PT: prothrombin time

According to WGCNA, there were five modules (clusters) among the 776 metabolites, which we named (by color) Green, Turquoise, Brown, Blue, and Yellow (**Figure 1**). A sixth module, the Grey module, included all metabolites that did not belong to any of the five modules. Compositions of the metabolite classifications (for example, lipids or amino acids) for these modules can be found in Figure S1; membership details for each module are listed in **Table S1**. Correlations between coagulation markers and eigenmetabolites are shown in **Figure 2**; PT was significantly and positively associated with the eigenmetabolite of the Green module, whose member metabolites belonged exclusively to the lipid super pathway (**Table S1**).

**Figure 1.**
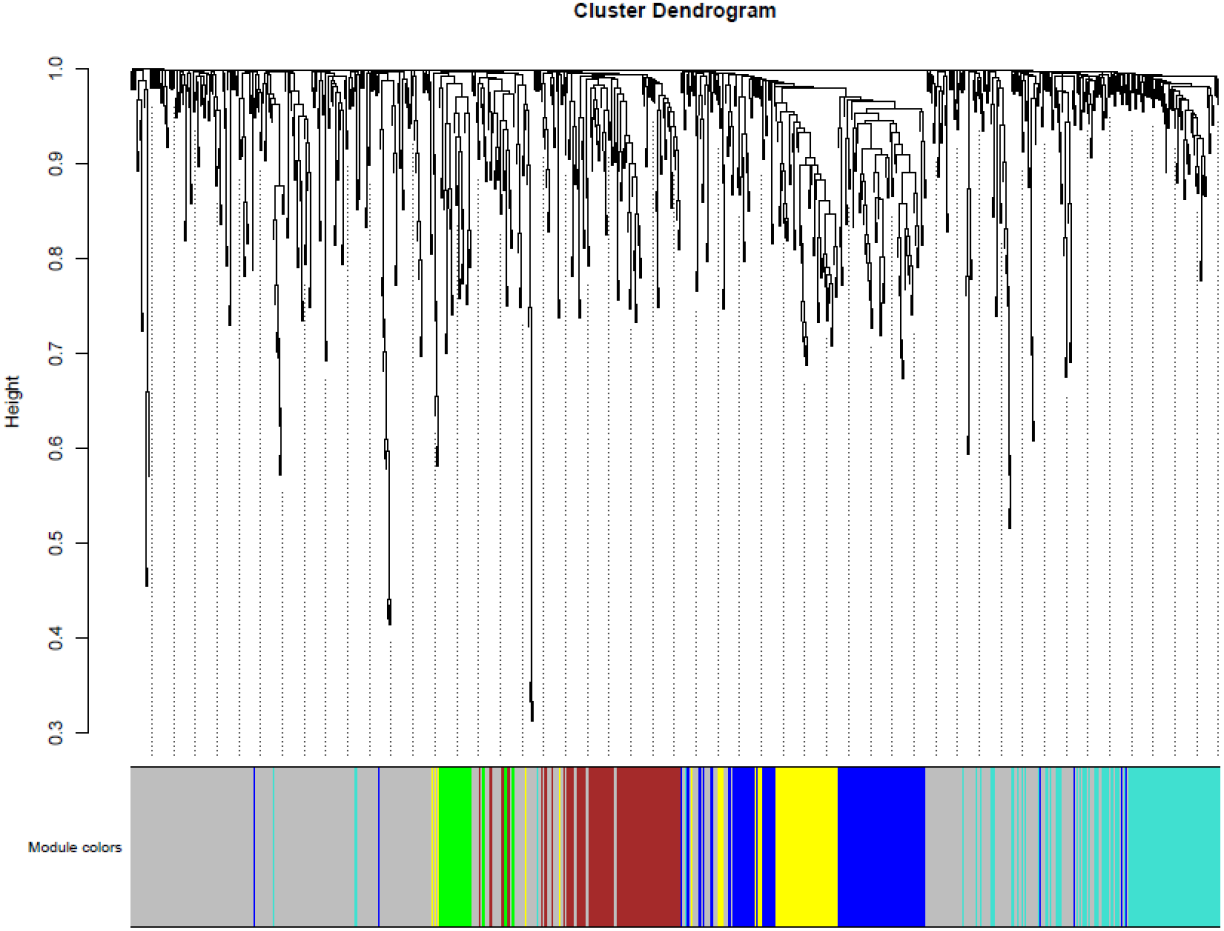
Metabolomic modules according to WGCNA results. The y-axis represents relative hierarchical distance across submodules. WGCNA: weighted gene co-expression network analysis

**Figure 2.**
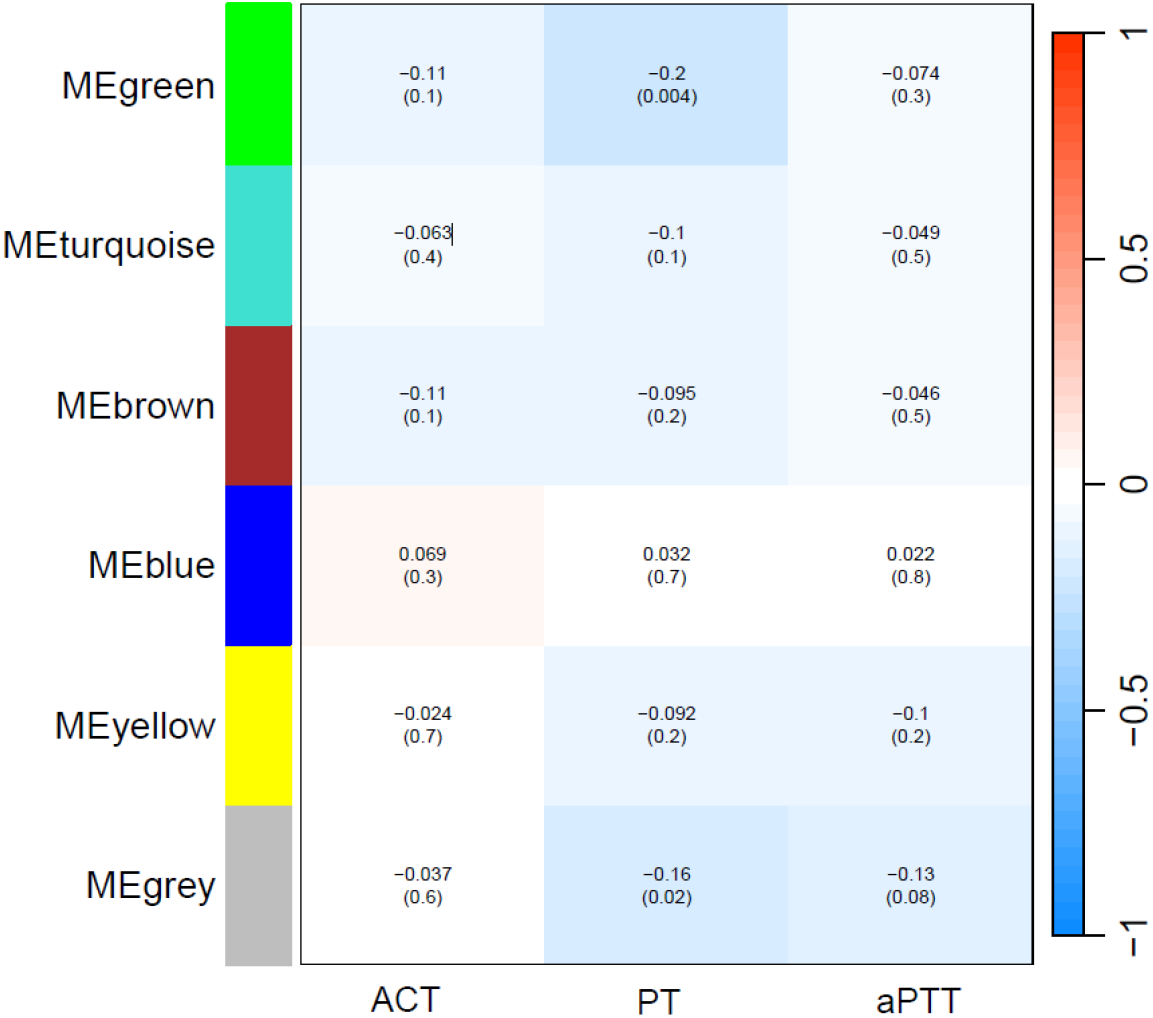
Correlations of metabolite modules with coagulation markers. Robust biweight midcorrelations are reported with p-values in parentheses. ACT: activated clotting time; aPTT: activated partial thromboplastin time; PT: prothrombin time

## Discussion

In the current study, we identified one metabolite (valine) associated with ACT and 20 associated with PT, with no metabolite associated with aPTT. We also demonstrated that metabolites in the blood tended to cluster into groups within which member metabolites were closely correlated. To our knowledge, this is among the first studies to systematically examine associations between the circulating metabolome and commonly used coagulation markers in young patients with CHD.

The number of metabolomic studies investigating coagulation markers in the literature remains limited. As the only metabolite associated with ACT, valine was a branched chain amino acid, the metabolism of which has been shown to promote thrombosis risk by enhancing tropomodulin-3 propionylation in platelets[13]. The observed inverse association between valine and ACT is broadly consistent with prior evidence linking branched-chain amino acid metabolism to thrombosis-related pathways. However, given the cross-sectional nature of the study, these findings should be interpreted as associations rather than evidence of causality. Several metabolites associated with PT are involved in lipid metabolism, including lysophosphatidylinositol species, sphingomyelins, a plasmalogen phosphatidylcholine species, and oleoyl-arachidonoyl-glycerol, which are involved in membrane organization and signaling processes related to platelet activation and coagulation[14,15]. In addition, the associations of retinol, hyocholate, biliverdin, and bilirubin degradation products with PT point to a potential role of hepatic and hepatobiliary metabolism, which is closely linked to coagulation factor synthesis and maintenance of hemostatic balance[16,17]. It is worth noting that retinol has been shown to induce platelet aggregation via activation of phospholipase A2[18]. Collectively, these associations suggest that variation in PT may reflect multiple biological processes, including lipid metabolism, hepatobiliary function, and pathways involved in platelet activation and coagulation. Nevertheless, mechanistic studies will be required to determine whether these metabolites directly influence hemostatic processes or serve as biomarkers of broader physiological states.

Clustering of metabolites from the WGCNA analysis indicates, as expected, that metabolites in the body are closely correlated. Notably, the Green module showed a significant correlation with PT. The Green module was composed predominantly of phosphatidylethanolamine, phosphatidylcholine, phosphatidylinositol, and diacylglycerol species, many of which contained arachidonic acid, linoleic acid, or docosahexaenoic acid side chains. These lipid classes are key components of membrane remodeling and lipid signaling pathways that have been implicated in platelet activation and thrombin generation through the production of bioactive lipid mediators and regulation of membrane-dependent coagulation processes[14,19]. The coordinated clustering of these metabolites suggests that phospholipid metabolism may contribute to interindividual variation in PT and potentially reflect alterations in platelet-dependent hemostatic pathways. The borderline correlation between the Grey module and PT was likely due to chance, as the module was mostly composed of unrelated metabolites.

The current study has strengths and limitations. We adopted rigorous quality assurance and control protocols, including consistency in the timing of blood draws and processing of blood samples and the use of blind duplicate blood samples. As for limitations, the analyses were cross-sectional in nature, and the associations observed in the study might be subject to potential bias. The coagulation markers were retrieved from the electronic medical record and were not measured for research purposes. Some coagulation markers were measured at different days than when the blood was drawn for metabolomic profiling. Blood samples were collected from a central line, which is a known source of heparin contamination. Accordingly, residual measurement error and unmeasured confounding cannot be excluded. These factors may have attenuated or inflated some observed associations. Further, children with CHD in the study had complex medical conditions, which could not be fully accounted for in statistical analyses. An independent validation cohort was not available because of the specialized nature of the study population. Consequently, the findings should be considered exploratory until replicated externally. Finally, our study sample was from young patients with CHD who needed corrective surgery, which limits the generalizability of our findings.

## 5. Conclusions

In conclusion, we identified 21 metabolites associated with PT or ACT in children with CHD undergoing cardiac surgery. These findings provide preliminary insights into potential metabolic correlates of coagulation markers in pediatric cardiovascular disease. Independent replication and mechanistic studies are needed before clinical implications can be established.

## Author Contributions

Conceptualization, S.L., D.W., and M.T.H.; methodology, S.L. and D.W.; formal analysis, S.L. and D.W.; investigation, S.L., D.W., A.J., Z.A., K.G., B.D., L.Z., W.T., D.M.O., and M.T.H; resources, S.L. D.M.O, and M.T.H.; data curation, A.J., Z.A., K.G. and M.T.H; writing—original draft preparation, S.L.; writing—review and editing, D.W., A.J., Z.A., K.G., B.D., L.Z., W.T., D.M.O., and M.T.H.; supervision, S.L. and M.T.H.; project administration, S.L., A.J., Z.A., and M.T.H; resources, S.L. D.M.O., and M.T.H.; funding acquisition, S.L. and M.T.H. All authors have read and agreed to the published version of the manuscript.

## Institutional Review Board Statement

The current study was approved by the Institutional Review Board of Children’s Minnesota.

## Informed Consent Statement

Parents or legal guardians provided written informed consent. Children 7-years or older also provided written assent when able.

## Data Availability Statement

The data that support the findings of this study are available from the corresponding author, upon reasonable request.

## Acknowledgments

The current study involved many colleagues at Children’s Minnesota, including colleagues at the Minneapolis Central Lab, the Cardiovascular Care Center, the Children’s Heart Clinic, and anesthesiologists, whose contributions are greatly appreciated. The authors of the study also wish to thank the children who participated in the study and their families. The authors thank Martin Cozza for his help with editing the manuscript.

