## Supplementary material for "Circulating Metabolites Associated with Prothrombin Time and Activated Clotting Time in Children with Congenital Heart Disease: An Untargeted Metabolomics Study": Figure S1 and Table S1

Correspondence:

Shengxu Li, MD, PhD, MPH

And

Marnie T. Huntley, MD

The Children's Heart Clinic

2530 Chicago Avenue South, Suite 500

Minneapolis, MN 55404

### Figure S1

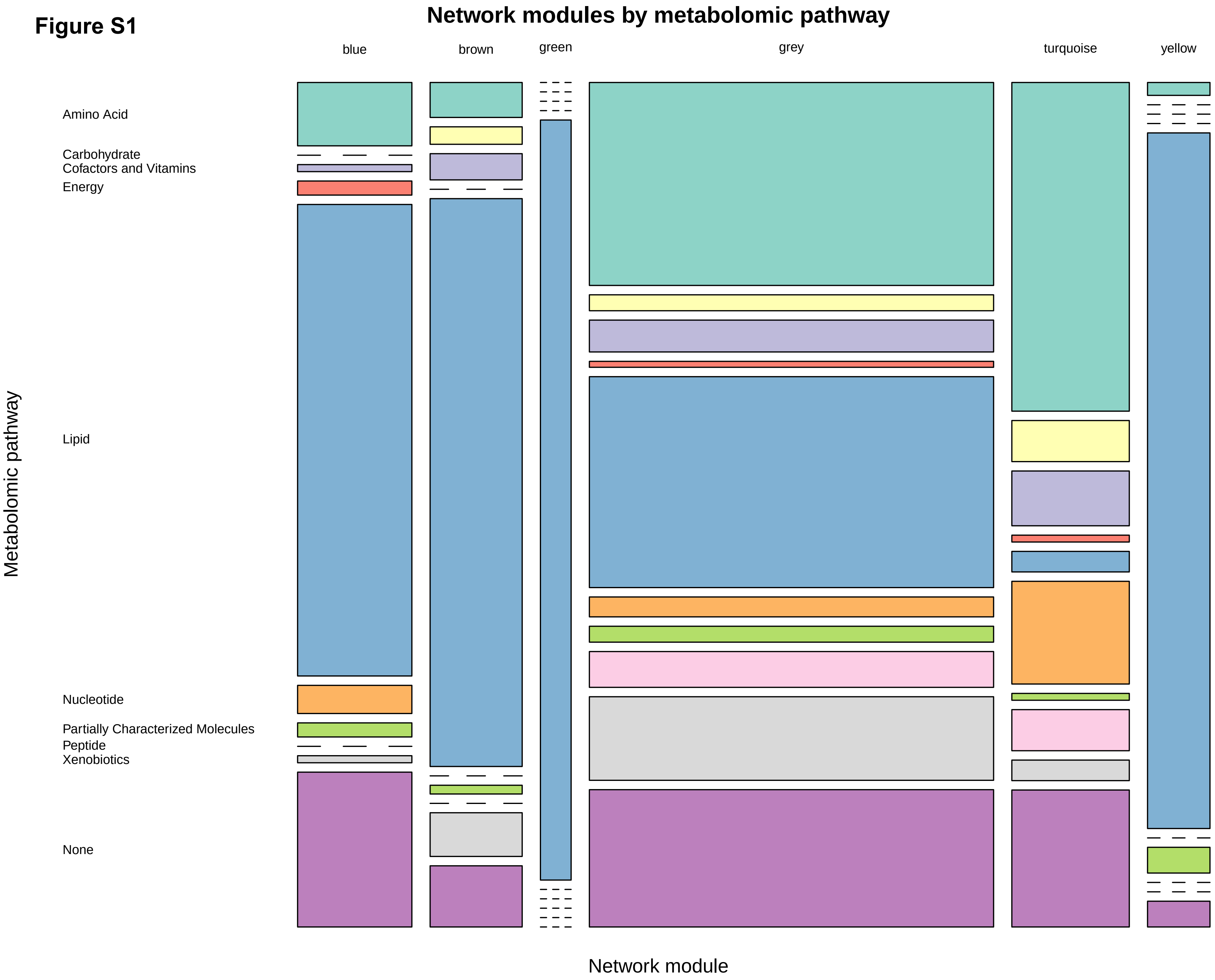
Network modules by metabolomic pathway
blue
brown
green
grey
turquoise
yellow
Amino Acid
Carbohydrate
Cofactors and Vitamins
Energy
Lipid
Nucleotide
Partially Characterized Molecules
Peptide
Xenobiotics
None
Metabolomic pathway
Network module

Table S1. Memberships of the modules from WGCNA

| Module Color | SUPER PATHWAY | SUB PATHWAY | CHEMICAL NAME | HMDB |
|---|---|---|---|---|
| blue | Lipid | Mevalonate Metabolism | 3-hydroxy-3-methylglutarate | HMDB0000355 |
| blue | Nucleotide | Purine Metabolism, Adenine containing | adenosine 3',5'-cyclic monophosphate (cAMP) | HMDB0000058 |
| blue | Lipid | Ketone Bodies | 3-hydroxybutyrate (BHBA) | HMDB0000442,HMDB0000357,HMDB0000011 |
| blue | Nucleotide | Pyrimidine Metabolism, Orotate containing | orotate | HMDB0000226 |
| blue | Amino Acid | Methionine, Cysteine, SAM and Taurine Metabolism | taurine | HMDB0000251 |
| blue | Amino Acid | Glycine, Serine and Threonine Metabolism | betaine | HMDB0000043 |
| blue | Amino Acid | Glycine, Serine and Threonine Metabolism | dimethylglycine | HMDB0000092 |
| blue | Nucleotide | Purine Metabolism, (Hypo)Xanthine/Inosine containing | xanthine | HMDB0000292 |
| blue | Nucleotide | Pyrimidine Metabolism, Thymine containing | 3-aminoisobutyrate | HMDB0002166 |
| blue | Energy | TCA Cycle | citrate | HMDB0000094 |
| blue | Amino Acid | Tryptophan Metabolism | anthranilate | HMDB0001123 |
| blue | Amino Acid | Leucine, Isoleucine and Valine Metabolism | methylsuccinate | HMDB0001844 |
| blue | Amino Acid | Leucine, Isoleucine and Valine Metabolism | ethylmalonate | HMDB0000622 |
| blue | Lipid | Fatty Acid, Dicarboxylate | suberate (C8-DC) | HMDB0000893 |
| blue | Lipid | Fatty Acid, Monohydroxy | 3-hydroxyoctanoate | HMDB0001954 |
| blue | Lipid | Fatty Acid Metabolism (Acyl Carnitine, Long Chain Saturated) | palmitoylcarnitine (C16) | HMDB0000222 |
| blue | Lipid | Fatty Acid Metabolism (Acyl Carnitine, Medium Chain) | hexanoylcarnitine (C6) | HMDB0000756 |
| blue | Lipid | Fatty Acid Metabolism (Acyl Carnitine, Short Chain) | acetylcarnitine (C2) | HMDB0000201 |
| blue | Lipid | Fatty Acid Metabolism (also BCAA Metabolism) | butyrylcarnitine (C4) | HMDB0002013 |
| blue | Lipid | Fatty Acid, Dicarboxylate | dodecanedioate (C12-DC) | HMDB0000623 |
| blue | Lipid | Fatty Acid, Monohydroxy | 3-hydroxysebacate | HMDB0000350 |
| blue | Lipid | Fatty Acid, Monohydroxy | 5-hydroxyhexanoate | HMDB0000409,HMDB0000525 |
| blue | Lipid | Fatty Acid, Dicarboxylate | sebacate (C10-DC) | HMDB0000792 |
| blue | Lipid | Fatty Acid Metabolism (Acyl Carnitine, Medium Chain) | octanoylcarnitine (C8) | HMDB0000791 |
| blue | Lipid | Fatty Acid Metabolism (Acyl Carnitine, Medium Chain) | decanoylcarnitine (C10) | HMDB0000651 |
| blue | Lipid | Fatty Acid Metabolism (Acyl Carnitine, Long Chain Saturated) | myristoylcarnitine (C14) | HMDB0005066 |
| blue | Energy | TCA Cycle | aconitate [cis or trans] | HMDB0000958,HMDB000072 |
| blue | Lipid | Fatty Acid Metabolism (Acyl Carnitine, Long Chain Saturated) | stearoylcarnitine (C18) | HMDB0000848 |
| blue | Lipid | Fatty Acid Metabolism (Acyl Carnitine, Medium Chain) | laurylcarnitine (C12) | HMDB000225 |
| blue | Lipid | Fatty Acid, Dicarboxylate | 3-hydroxydodecanedioate* | HMDB0000413 |
| blue | Lipid | Fatty Acid Metabolism (Acyl Carnitine, Monounsaturated) | oleoylcarnitine (C18:1) | HMDB0005065 |
| blue | Lipid | Fatty Acid, Dicarboxylate | tetradecanedioate (C14-DC) | HMDB0000872 |
| blue | Lipid | Fatty Acid, Dicarboxylate | hexadecanedioate (C16-DC) | HMDB0000672 |
| blue | Lipid | Fatty Acid, Dicarboxylate | octadecanedioate (C18-DC) | HMDB0000782 |
| blue | Lipid | Carnitine Metabolism | deoxycarnitine | HMDB0001161 |
| blue | Lipid | Fatty Acid Metabolism (Acyl Glutamine) | hexanoylglutamine | |
| blue | Cofactors and Vitamins | Hemoglobin and Porphyrin Metabolism | bilirubin (E,E)* | HMDB0240584 |
| blue | Lipid | Secondary Bile Acid Metabolism | taurocholenate sulfate* | |
| blue | Xenobiotics | Food Component/Plant | cinnamoylglycine | HMDB0011621 |
| blue | Lipid | Fatty Acid Metabolism (Acyl Carnitine, Monounsaturated) | cis-4-decenoylcarnitine (C10:1) | HMDB0013205 |
| blue | Lipid | Fatty Acid, Dihydroxy | 2S,3R-dihydroxybutyrate | HMDB0002453 |
| blue | Lipid | Fatty Acid Metabolism (Acyl Carnitine, Polyunsaturated) | linoleoylcarnitine (C18:2)* | HMDB0006469 |
| blue | Lipid | Fatty Acid Metabolism (Acyl Carnitine, Hydroxy) | (R)-3-hydroxybutyrylcarnitine | HMDB0013127 |
| blue | Lipid | Fatty Acid Metabolism (Acyl Carnitine, Dicarboxylate) | octadecenedioylcarnitine (C18:1-DC)* | |
| blue | Lipid | Fatty Acid Metabolism (Acyl Carnitine, Dicarboxylate) | octadecanedioylcarnitine (C18-DC)* | |
| blue | Lipid | Fatty Acid Metabolism (Acyl Carnitine, Monounsaturated) | myristoleoylcarnitine (C14:1)* | HMDB0240588 |
| blue | Lipid | Fatty Acid, Monohydroxy | 3-hydroxyhexanoate | HMDB0061652,HMDB0010718 |
| blue | Lipid | Fatty Acid Metabolism (Acyl Carnitine, Dicarboxylate) | adipoylcarnitine (C6-DC) | HMDB0061677 |
| blue | Lipid | Fatty Acid Metabolism (Acyl Carnitine, Dicarboxylate) | suberoylcarnitine (C8-DC) | |
| blue | Amino Acid | Glutathione Metabolism | 2-hydroxybutyrate/2-hydroxyisobutyrate | HMDB0000729,HMDB00008 |
| blue | Lipid | Fatty Acid Metabolism (Acyl Carnitine, Hydroxy) | (S)-3-hydroxybutyrylcarnitine | HMDB0013127 |
| blue | Lipid | Fatty Acid Metabolism (Acyl Carnitine, Monounsaturated) | palmitoleoylcarnitine (C16:1)* | HMDB0013207 |
| blue | Lipid | Fatty Acid Metabolism (Acyl Carnitine, Dicarboxylate) | pimeloylcarnitine/3-methyladipoylcarnitine (C7-DC) | |
| blue | Lipid | Fatty Acid, Monohydroxy | 2-hydroxynervonate* | |
| blue | Lipid | Endocannabinoid | N-oleoylserine | |
| blue | Lipid | Fatty Acid Metabolism (Acyl Carnitine, Polyunsaturated) | linolenoylcarnitine (C18:3)* | |
| blue | Lipid | Fatty Acid Metabolism (Acyl Carnitine, Long Chain Saturated) | behenoylcarnitine (C22)* | HMDB0062468 |
| blue | Lipid | Fatty Acid Metabolism (Acyl Carnitine, Long Chain Saturated) | arachidoylcarnitine (C20)* | HMDB0006460 |
| blue | Lipid | Fatty Acid Metabolism (Acyl Carnitine, Long Chain Saturated) | lignoceroylcarnitine (C24)* | HMDB0240665 |
| blue | Lipid | Fatty Acid Metabolism (Acyl Carnitine, Long Chain Saturated) | cerotoylcarnitine (C26)* | HMDB0006347 |
| blue | Lipid | Fatty Acid Metabolism (Acyl Carnitine, Monounsaturated) | ximenoylcarnitine (C26:1)* | |
| blue | Lipid | Fatty Acid Metabolism (Acyl Carnitine, Polyunsaturated) | arachidonoylcarnitine (C20:4) | HMDB0006455 |
| blue | Lipid | Fatty Acid Metabolism (Acyl Carnitine, Monounsaturated) | eicosenoylcarnitine (C20:1)* | |
| blue | Lipid | Fatty Acid Metabolism (Acyl Carnitine, Polyunsaturated) | dihomo-linoleoylcarnitine (C20:2)* | |
| blue | Lipid | Fatty Acid Metabolism (Acyl Carnitine, Monounsaturated) | nervonoylcarnitine (C24:1)* | HMDB0006509 |
| blue | Lipid | Fatty Acid Metabolism (Acyl Carnitine, Monounsaturated) | 5-dodecenoylcarnitine (C12:1) | HMDB13326 |
| blue | Lipid | Fatty Acid Metabolism (Acyl Carnitine, Hydroxy) | 3-hydroxyoleoylcarnitine | |
| blue | Lipid | Fatty Acid Metabolism (Acyl Glycine) | trans-2-hexenoylglycine | |
| blue | Lipid | Fatty Acid, Dicarboxylate | dodecenedioate (C12:1-DC)* | HMDB0000933 |
| blue | Lipid | Fatty Acid, Dicarboxylate | hexadecenedioate (C16:1-DC)* | |
| blue | Lipid | Fatty Acid, Dicarboxylate | octadecenedioate (C18:1-DC) | |
| blue | Lipid | Fatty Acid, Dicarboxylate | octadecadienedioate (C18:2-DC)* | |
| blue | Lipid | Fatty Acid Metabolism (Acyl Glycine) | 3-hydroxybutyroylglycine** | |
| blue | Partially Characterized Molecules | Partially Characterized Molecules | glutamine conjugate of C6H10O2 (1)* | |
| blue | Partially Characterized Molecules | Partially Characterized Molecules | glutamine conjugate of C6H10O2 (2)* | |
| blue | Amino Acid | Tryptophan Metabolism | 8-methoxykynurenate | HMDB0060426 |
| blue | Lipid | Fatty Acid, Dicarboxylate | 2-hydroxysebacate | HMDB0000424 |
| blue | Lipid | Fatty Acid Metabolism (Acyl Carnitine, Hydroxy) | 3-hydroxyhexanoylcarnitine (1) | |
| blue | Lipid | Fatty Acid, Dicarboxylate | tetradecadienedioate (C14:2-DC)* | |
| blue | Lipid | Fatty Acid Metabolism (Acyl Carnitine, Hydroxy) | 3-hydroxydecanoylcarnitine | HMDB0061636 |
| blue | Amino Acid | Methionine, Cysteine, SAM and Taurine Metabolism | succinoyltaurine | |
| blue | Lipid | Fatty Acid, Dicarboxylate | decadienedioic acid (C10:2-DC)** | |
| blue | Lipid | Fatty Acid, Branched | cis-3,4-methyleneheptanoate | |
| blue | Lipid | Fatty Acid Metabolism (Acyl Carnitine, Medium Chain) | cis-3,4-methyleneheptanoylcarnitine | |
| blue | Lipid | Fatty Acid Metabolism (Acyl Carnitine, Hydroxy) | 3-hydroxyoctanoylcarnitine (1) | |
| blue | Lipid | Fatty Acid Metabolism (Acyl Carnitine, Hydroxy) | 3-hydroxyoctanoylcarnitine (2) | |
| blue | | | X-11478 | |
| blue | | | X-12101 | |
| blue | | | X-12839 | |
| blue | | | X-14939 | |
| blue | | | X-15486 | |
| blue | | | X-16397 | |
| blue | | | X-16580 | |
| blue | | | X-17335 | |
| blue | | | X-17438 | |
| blue | | | X-18886 | |
| blue | | | X-18888 | |
| blue | | | X-18913 | |
| blue | | | X-18921 | |
| blue | | | X-21364 | |
| blue | | | X-21736 | |
| blue | | | X-23665 | |
| blue | | | X-23678 | |
| blue | | | X-23680 | |
| blue | | | X-24949 | |
| blue | | | X-26106 | |
| blue | | | X-26107 | |
| blue | | | X-26108 | |
| brown | Lipid | Sterol | cholesterol | HMDB0000067 |
| brown | Lipid | Dihydroceramides | N-stearoyl-sphinganine (d18:0/18:0)* | HMDB0011761 |
| brown | Lipid | Phosphatidylcholine (PC) | 1-palmitoyl-2-linoleoyl-GPC (16:0/18:2) | HMDB0007973 |

| | | | | |
|---|---|---|---|---|
| brown | Lipid | Sphingomyelins | stearoyl sphingomyelin (d18:1/18:0) | HMDB0001348 |
| brown | Lipid | Phosphatidylcholine (PC) | 1-palmitoyl-2-oleoyl-GPC (16:0/18:1) | HMDB0007972 |
| brown | Lipid | Ceramides | N-stearoyl-sphingosine (d18:1/18:0)* | HMDB0004950 |
| brown | Carbohydrate | Glycolysis, Gluconeogenesis, and Pyruvate Metabolism | 1,5-anhydroglucitol (1,5-AG) | HMDB0002712 |
| brown | Lipid | Phosphatidylcholine (PC) | 1,2-dipalmitoyl-GPC (16:0/16:0) | HMDB0000564 |
| brown | Lipid | Phosphatidylcholine (PC) | 1-myristoyl-2-palmitoyl-GPC (14:0/16:0) | HMDB0007869 |
| brown | Carbohydrate | Fructose, Mannose and Galactose Metabolism | galactonate | HMDB0000565 |
| brown | Lipid | Medium Chain Fatty Acid | 10-undecenoate (11:1n1) | HMDB0033724 |
| brown | Lipid | Lysophospholipid | 1-palmitoyl-GPC (16:0) | HMDB0010382 |
| brown | Xenobiotics | Food Component/Plant | homostachydrine* | HMDB0033433 |
| brown | Amino Acid | Tryptophan Metabolism | tryptophan betaine | HMDB0061115 |
| brown | Lipid | Hexosylceramides (HCER) | glycosyl-N-stearoyl-sphingosine (d18:1/18:0) | |
| brown | Cofactors and Vitamins | Vitamin A Metabolism | beta-cryptoxanthin | HMDB0033844 |
| brown | Lipid | Sphingomyelins | sphingomyelin (d18:1/18:1, d18:2/18:0) | HMDB0012101 |
| brown | Lipid | Sphingomyelins | palmitoyl sphingomyelin (d18:1/16:0) | HMDB0010169 |
| brown | Xenobiotics | Food Component/Plant | ergothioneine | HMDB0003045 |
| brown | Lipid | Fatty Acid, Dicarboxylate | tridecenedioate (C13:1-DC)* | |
| brown | Amino Acid | Methionine, Cysteine, SAM and Taurine Metabolism | S-methylcysteine | HMDB0002108 |
| brown | Lipid | Lysoplasmalogen | 1-(1-enyl-palmitoyl)-GPC (P-16:0)* | HMDB0010407 |
| brown | Lipid | Lysoplasmalogen | 1-(1-enyl-palmitoyl)-GPE (P-16:0)* | HMDB0011152 |
| brown | Lipid | Lysoplasmalogen | 1-(1-enyl-stearoyl)-GPE (P-18:0)* | HMDB0240598 |
| brown | Lipid | Fatty Acid Metabolism (Acyl Carnitine, Long Chain Saturated) | margaroylcarnitine (C17)* | HMDB0006210 |
| brown | Lipid | Sphingomyelins | sphingomyelin (d18:1/14:0, d16:1/16:0)* | HMDB0012097 |
| brown | Lipid | Sphingomyelins | sphingomyelin (d18:2/16:0, d18:1/16:1)* | HMDB0240638,HMDB0240613 |
| brown | Lipid | Sphingomyelins | sphingomyelin (d18:2/14:0, d18:1/14:1)* | HMDB0240637,HMDB0240612 |
| brown | Lipid | Sphingomyelins | sphingomyelin (d18:1/24:1, d18:2/24:0)* | HMDB0012107 |
| brown | Xenobiotics | Chemical | 4-hydroxychlorothalonil | HMDB0240624 |
| brown | Lipid | Sphingomyelins | sphingomyelin (d18:1/20:0, d16:1/22:0)* | HMDB0012102 |
| brown | Lipid | Sphingomyelins | sphingomyelin (d18:1/20:1, d18:2/20:0)* | HMDB0240610,HMDB0240632 |
| brown | Lipid | Sphingomyelins | behenoyl sphingomyelin (d18:1/22:0)* | HMDB0012103 |
| brown | Lipid | Sphingomyelins | sphingomyelin (d18:1/22:2, d18:2/22:1, d16:1/24:2)* | HMDB0240670,HMDB0240672,HMDB0240669 |
| brown | Lipid | Sphingomyelins | lignoceroyl sphingomyelin (d18:1/24:0) | HMDB0011697 |
| brown | Lipid | Sphingomyelins | sphingomyelin (d17:1/16:0, d18:1/15:0, d16:1/17:0)* | HMDB0240617,HMDB0240608 |
| brown | Lipid | Plasmalogen | 1-(1-enyl-stearoyl)-2-oleoyl-GPE (P-18:0/18:1) | HMDB0011375 |
| brown | Lipid | Sphingomyelins | sphingomyelin (d18:1/17:0, d17:1/18:0, d19:1/16:0) | HMDB0240620,HMDB0240609,HMDB0240622 |
| brown | Lipid | Phosphatidylcholine (PC) | 1-palmitoyl-2-stearoyl-GPC (16:0/18:0) | HMDB0007970 |
| brown | Lipid | Dihydrosphingomyelins | palmitoyl dihydrosphingomyelin (d18:0/16:0)* | HMDB0010168 |
| brown | Lipid | Sphingomyelins | tricosanoyl sphingomyelin (d18:1/23:0)* | HMDB0012105 |
| brown | Lipid | Sphingomyelins | sphingomyelin (d18:2/23:0, d18:1/23:1, d17:1/24:1)* | HMDB0240634,HMDB0011696,HMDB0240614 |
| brown | Lipid | Phosphatidylcholine (PC) | 1-palmitoyl-2-palmitoleoyl-GPC (16:0/16:1)* | HMDB0007969 |
| brown | Lipid | Plasmalogen | 1-(1-enyl-stearoyl)-2-arachidonoyl-GPE (P-18:0/20:4)* | HMDB0005779 |
| brown | Lipid | Plasmalogen | 1-(1-enyl-palmitoyl)-2-arachidonoyl-GPE (P-16:0/20:4)* | HMDB0011352 |
| brown | Lipid | Plasmalogen | 1-(1-enyl-palmitoyl)-2-arachidonoyl-GPC (P-16:0/20:4)* | HMDB0011220 |
| brown | Lipid | Sphingomyelins | sphingomyelin (d18:1/21:0, d17:1/22:0, d16:1/23:0)* | HMDB0240619,HMDB0240611,HMDB0240621 |
| brown | Lipid | Dihydrosphingomyelins | behenoyl dihydrosphingomyelin (d18:0/22:0)* | HMDB0012091 |
| brown | Lipid | Dihydrosphingomyelins | sphingomyelin (d18:0/18:0, d19:0/17:0)* | HMDB0012087 |
| brown | Lipid | Dihydroceramides | N-palmitoyl-sphinganine (d18:0/16:0) | HMDB11760 |
| brown | Lipid | Dihydrosphingomyelins | myristoyl dihydrosphingomyelin (d18:0/14:0)* | HMDB0012085 |
| brown | Lipid | Plasmalogen | 1-(1-enyl-palmitoyl)-2-palmitoyl-GPC (P-16:0/16:0)* | HMDB0011206 |
| brown | Lipid | Plasmalogen | 1-(1-enyl-stearoyl)-2-linoleoyl-GPE (P-18:0/18:2)* | HMDB0011376 |
| brown | Lipid | Hexosylceramides (HCER) | glycosyl-N-palmitoyl-sphingosine (d18:1/16:0) | |
| brown | Lipid | Lactosylceramides (LCER) | lactosyl-N-behenoyl-sphingosine (d18:1/22:0)* | HMDB0011594 |
| brown | Lipid | Hexosylceramides (HCER) | glycosyl-N-behenoyl-sphingadienine (d18:2/22:0)* | |
| brown | Lipid | Ceramides | ceramide (d16:1/24:1, d18:1/22:1)* | |
| brown | Lipid | Ceramides | N-palmitoyl-heptadecasphingosine (d17:1/16:0)* | HMDB0240685 |
| brown | Lipid | Ceramides | ceramide (d18:2/24:1, d18:1/24:2)* | HMDB0240679,HMDB0240680 |
| brown | Lipid | Hexosylceramides (HCER) | glycosyl ceramide (d18:2/24:1, d18:1/24:2)* | |
| brown | Lipid | Hexosylceramides (HCER) | glycosyl-N-tricosanoyl-sphingadienine (d18:2/23:0)* | |
| brown | Lipid | Hexosylceramides (HCER) | glycosyl ceramide (d18:1/23:1, d17:1/24:1)* | |
| brown | Lipid | Dihydrosphingomyelins | sphingomyelin (d18:0/20:0, d16:0/22:0)* | HMDB0012090 |
| brown | Lipid | Sphingomyelins | sphingomyelin (d18:1/19:0, d19:1/18:0)* | |
| brown | Lipid | Sphingomyelins | sphingomyelin (d18:2/18:1)* | HMDB0001348 |
| brown | Lipid | Sphingomyelins | sphingomyelin (d18:2/21:0, d16:2/23:0)* | HMDB0240676 |
| brown | Lipid | Sphingomyelins | sphingomyelin (d18:2/23:1)* | HMDB0240668 |
| brown | Lipid | Sphingomyelins | sphingomyelin (d18:1/25:0, d19:0/24:1, d20:1/23:0, d19:1/24:0)* | HMDB0240675,HMDB0240674,HMDB0240673,HMDB0240671 |
| brown | Lipid | Sphingomyelins | sphingomyelin (d17:2/16:0, d18:2/15:0)* | HMDB0240677 |
| brown | Lipid | Hexosylceramides (HCER) | glycosyl ceramide (d18:1/20:0, d16:1/22:0)* | |
| brown | Amino Acid | Lysine Metabolism | N,N,N-trimethyl-5-aminovalerate | |
| brown | Cofactors and Vitamins | Vitamin A Metabolism | carotene diol (1) | |
| brown | Cofactors and Vitamins | Vitamin A Metabolism | carotene diol (3) | |
| brown | Partially Characterized Molecules | Partially Characterized Molecules | glutamine conjugate of C7H12O2* | |
| brown | Lipid | Sphingomyelins | sphingomyelin (d17:1/14:0, d16:1/15:0)* | |
| brown | Amino Acid | Tryptophan Metabolism | 6-bromotryptophan | |
| brown | Lipid | Fatty Acid Metabolism (Acyl Carnitine, Monounsaturated) | undecenoylcarnitine (C11:1) | |
| brown | Lipid | Ceramide PEs | palmitoyl-sphingosine-phosphoethanolamine (d18:1/16:0) | |
| brown | Xenobiotics | Chemical | 3,5-dichloro-2,6-dihydroxybenzoic acid | |
| brown | Xenobiotics | Chemical | 3-bromo-5-chloro-2,6-dihydroxybenzoic acid* | |
| brown | | | X-11381 | |
| brown | | | X-11795 | |
| brown | | | X-12798 | |
| brown | | | X-18901 | |
| brown | | | X-24522 | |
| brown | | | X-24812 | |
| brown | | | X-25420 | |
| green | Lipid | Phosphatidylethanolamine (PE) | 1-palmitoyl-2-oleoyl-GPE (16:0/18:1) | HMDB0005320 |
| green | Lipid | Phosphatidylinositol (PI) | 1-palmitoyl-2-linoleoyl-GPI (16:0/18:2) | HMDB0009784 |
| green | Lipid | Phosphatidylinositol (PI) | 1-stearoyl-2-arachidonoyl-GPI (18:0/20:4) | HMDB0009815 |
| green | Lipid | Lysophospholipid | 1-stearoyl-GPE (18:0) | HMDB0011130 |
| green | Lipid | Lysophospholipid | 1-arachidonoyl-GPC (20:4n6)* | HMDB0010395 |
| green | Lipid | Lysophospholipid | 1-palmitoyl-GPE (16:0) | HMDB0011503 |
| green | Lipid | Lysophospholipid | 1-arachidonoyl-GPE (20:4n6)* | HMDB0011517 |
| green | Lipid | Phosphatidylethanolamine (PE) | 1-stearoyl-2-oleoyl-GPE (18:0/18:1) | HMDB0008993 |
| green | Lipid | Phosphatidylcholine (PC) | 1-stearoyl-2-arachidonoyl-GPC (18:0/20:4) | HMDB0008048 |
| green | Lipid | Phosphatidylethanolamine (PE) | 1-palmitoyl-2-linoleoyl-GPE (16:0/18:2) | HMDB0005322 |
| green | Lipid | Diacylglycerol | oleoyl-linoleoyl-glycerol (18:1/18:2) [1] | HMDB0007219 |
| green | Lipid | Diacylglycerol | oleoyl-linoleoyl-glycerol (18:1/18:2) [2] | HMDB0007219 |
| green | Lipid | Phosphatidylcholine (PC) | 1-palmitoyl-2-arachidonoyl-GPC (16:0/20:4n6) | HMDB0007982 |
| green | Lipid | Phosphatidylethanolamine (PE) | 1-stearoyl-2-linoleoyl-GPE (18:0/18:2)* | HMDB0008994 |
| green | Lipid | Phosphatidylethanolamine (PE) | 1-stearoyl-2-arachidonoyl-GPE (18:0/20:4) | HMDB0009003 |
| green | Lipid | Phosphatidylethanolamine (PE) | 1-palmitoyl-2-arachidonoyl-GPE (16:0/20:4)* | HMDB0005323 |
| green | Lipid | Phosphatidylethanolamine (PE) | 1-palmitoyl-2-docosahexaenoyl-GPE (16:0/22:6)* | HMDB0008946 |
| green | Lipid | Phosphatidylinositol (PI) | 1-palmitoyl-2-arachidonoyl-GPI (16:0/20:4)* | HMDB0009789 |
| green | Lipid | Diacylglycerol | palmitoyl-linoleoyl-glycerol (16:0/18:2) [2]* | HMDB0007103 |
| green | Lipid | Diacylglycerol | oleoyl-oleoyl-glycerol (18:1/18:1) [1]* | HMDB0007218 |
| green | Lipid | Diacylglycerol | oleoyl-oleoyl-glycerol (18:1/18:1) [2]* | HMDB0007218 |
| green | Lipid | Diacylglycerol | linoleoyl-arachidonoyl-glycerol (18:2/20:4) [1]* | HMDB0007257 |
| green | Lipid | Diacylglycerol | linoleoyl-arachidonoyl-glycerol (18:2/20:4) [2]* | HMDB0007257 |
| green | Lipid | Diacylglycerol | linoleoyl-docosahexaenoyl-glycerol (18:2/22:6) [2]* | HMDB0007266 |
| green | Lipid | Diacylglycerol | oleoyl-arachidonoyl-glycerol (18:1/20:4) [1]* | HMDB0007228 |
| green | Lipid | Diacylglycerol | oleoyl-arachidonoyl-glycerol (18:1/20:4) [2]* | HMDB0007228 |
| green | Lipid | Diacylglycerol | diacylglycerol (16:1/18:2 [2], 16:0/18:3 [1])* | |
| green | Lipid | Diacylglycerol | linoleoyl-linoleoyl-glycerol (18:2/18:2) [1]* | HMDB0007248 |

| | | | | |
|---|---|---|---|---|
| green | Lipid | Diacylglycerol | linoleoyl-linoleoyl-glycerol (18:2/18:2) [2]* | HMDB0007248 |
| grey | Amino Acid | Polyamine Metabolism | spermidine | HMDB0001257 |
| grey | Cofactors and Vitamins | Nicotinate and Nicotinamide Metabolism | 1-methylnicotinamide | HMDB0000699 |
| grey | Lipid | Fatty Acid, Dihydroxy | 12,13-DiHOME | HMDB0004705 |
| grey | Energy | TCA Cycle | alpha-ketoglutarate | HMDB0000208 |
| grey | Amino Acid | Leucine, Isoleucine and Valine Metabolism | 3-hydroxyisobutyrate | HMDB0000023,HMDB0000336 |
| grey | Carbohydrate | Glycolysis, Gluconeogenesis, and Pyruvate Metabolism | 3-phosphoglycerate | HMDB0000807 |
| grey | Lipid | Primary Bile Acid Metabolism | cholate | HMDB0000619 |
| grey | Nucleotide | Purine Metabolism, (Hypo)Xanthine/Inosine containing | hypoxanthine | HMDB0000157 |
| grey | Lipid | Fatty Acid, Dihydroxy | 9,10-DiHOME | HMDB0004704 |
| grey | Nucleotide | Purine Metabolism, Adenine containing | adenosine 5'-monophosphate (AMP) | HMDB0000045 |
| grey | Nucleotide | Purine Metabolism, Adenine containing | N6-methyladenosine | HMDB0004044 |
| grey | Amino Acid | Urea cycle; Arginine and Proline Metabolism | arginine | HMDB0000517 |
| grey | Amino Acid | Alanine and Aspartate Metabolism | aspartate | HMDB0000191 |
| grey | Amino Acid | Tyrosine Metabolism | 3-(4-hydroxyphenyl)lactate | HMDB0000755 |
| grey | Amino Acid | Phenylalanine Metabolism | phenylpyruvate | HMDB0000205 |
| grey | Amino Acid | Histidine Metabolism | carnosine | HMDB0000033 |
| grey | Cofactors and Vitamins | Hemoglobin and Porphyrin Metabolism | biliverdin | HMDB0001008 |
| grey | Lipid | Phospholipid Metabolism | choline phosphate | HMDB0001565 |
| grey | Lipid | Corticosteroids | cortisone | HMDB0002802 |
| grey | Amino Acid | Glutathione Metabolism | cysteinylglycine | HMDB0000078 |
| grey | Lipid | Sphingosines | sphingosine | HMDB0000252 |
| grey | Peptide | Gamma-glutamyl Amino Acid | gamma-glutamylglutamate | HMDB0011737 |
| grey | Xenobiotics | Food Component/Plant | gluconate | HMDB0000625 |
| grey | Amino Acid | Glycine, Serine and Threonine Metabolism | glycine | HMDB0000123 |
| grey | Lipid | Primary Bile Acid Metabolism | glycocholate | HMDB0000138 |
| grey | Amino Acid | Creatine Metabolism | guanidinoacetate | HMDB0000128 |
| grey | Amino Acid | Histidine Metabolism | histidine | HMDB0000177 |
| grey | Lipid | Corticosteroids | cortisol | HMDB0000063 |
| grey | Amino Acid | Methionine, Cysteine, SAM and Taurine Metabolism | hypotaurine | HMDB0000965 |
| grey | Nucleotide | Purine Metabolism, (Hypo)Xanthine/Inosine containing | inosine | HMDB0000195 |
| grey | Lipid | Inositol Metabolism | myo-inositol | HMDB0000211 |
| grey | Amino Acid | Leucine, Isoleucine and Valine Metabolism | isoleucine | HMDB0000172 |
| grey | Amino Acid | Lysine Metabolism | 2-aminoadipate | HMDB0000510 |
| grey | Amino Acid | Urea cycle; Arginine and Proline Metabolism | citrulline | HMDB0000904 |
| grey | Amino Acid | Leucine, Isoleucine and Valine Metabolism | leucine | HMDB0000687 |
| grey | Amino Acid | Lysine Metabolism | lysine | HMDB0003405 |
| grey | Energy | TCA Cycle | malate | HMDB0031518,HMDB0000156,HMDB0000744 |
| grey | Amino Acid | Methionine, Cysteine, SAM and Taurine Metabolism | methionine | HMDB0000696 |
| grey | Lipid | Fatty Acid Metabolism (also BCAA Metabolism) | methylmalonate (MMA) | HMDB0000202 |
| grey | Cofactors and Vitamins | Nicotinate and Nicotinamide Metabolism | nicotinamide | HMDB0001406 |
| grey | Lipid | Medium Chain Fatty Acid | pelargonate (9:0) | HMDB0000847 |
| grey | Amino Acid | Urea cycle; Arginine and Proline Metabolism | ornithine | HMDB0000214 |
| grey | Amino Acid | Phenylalanine Metabolism | phenylalanine | HMDB0000159 |
| grey | Energy | Oxidative Phosphorylation | phosphate | HMDB0001429 |
| grey | Xenobiotics | Food Component/Plant | phytanate | HMDB0000801 |
| grey | Amino Acid | Urea cycle; Arginine and Proline Metabolism | proline | HMDB0000162,HMDB0003411 |
| grey | Carbohydrate | Glycolysis, Gluconeogenesis, and Pyruvate Metabolism | lactate | HMDB0000190 |
| grey | Cofactors and Vitamins | Vitamin B6 Metabolism | pyridoxal | HMDB0001545 |
| grey | Xenobiotics | Drug - Topical Agents | salicylate | HMDB0001895 |
| grey | Amino Acid | Glycine, Serine and Threonine Metabolism | serine | HMDB0000187 |
| grey | Nucleotide | Pyrimidine Metabolism, Uracil containing | uridine | HMDB0000296 |
| grey | Nucleotide | Pyrimidine Metabolism, Uracil containing | 2'-deoxyuridine | HMDB0000012 |
| grey | Amino Acid | Histidine Metabolism | trans-urocanate | HMDB0000301 |
| grey | Amino Acid | Glutamate Metabolism | glutamate | HMDB0000148 |
| grey | Amino Acid | Glutamate Metabolism | glutamine | HMDB0000641 |
| grey | Amino Acid | Glycine, Serine and Threonine Metabolism | threonine | HMDB0000167 |
| grey | Amino Acid | Tryptophan Metabolism | tryptophan | HMDB0000929 |
| grey | Amino Acid | Leucine, Isoleucine and Valine Metabolism | valine | HMDB0000883 |
| grey | Carbohydrate | Glycolysis, Gluconeogenesis, and Pyruvate Metabolism | glucose | HMDB0000122 |
| grey | Carbohydrate | Fructose, Mannose and Galactose Metabolism | mannose | HMDB0000169 |
| grey | Amino Acid | Alanine and Aspartate Metabolism | alanine | HMDB0000161 |
| grey | Amino Acid | Tyrosine Metabolism | tyrosine | HMDB0000158 |
| grey | Carbohydrate | Glycolysis, Gluconeogenesis, and Pyruvate Metabolism | pyruvate | HMDB0000243 |
| grey | Nucleotide | Pyrimidine Metabolism, Thymine containing | thymidine | HMDB0000273 |
| grey | Amino Acid | Alanine and Aspartate Metabolism | asparagine | HMDB0000168 |
| grey | Lipid | Medium Chain Fatty Acid | caprylate (8:0) | HMDB0000482 |
| grey | Amino Acid | Urea cycle; Arginine and Proline Metabolism | trans-4-hydroxyproline | HMDB0000725 |
| grey | Amino Acid | Glutathione Metabolism | 5-oxoproline | HMDB0000267 |
| grey | Amino Acid | Tryptophan Metabolism | picolinate | HMDB0002243 |
| grey | Amino Acid | Glycine, Serine and Threonine Metabolism | sarcosine | HMDB0000271 |
| grey | Amino Acid | Lysine Metabolism | pipecolate | HMDB0000070 |
| grey | Lipid | Phospholipid Metabolism | phosphoethanolamine | HMDB0000224 |
| grey | Cofactors and Vitamins | Hemoglobin and Porphyrin Metabolism | bilirubin (Z,Z) | HMDB0000054 |
| grey | Amino Acid | Tyrosine Metabolism | thyroxine | HMDB0000248 |
| grey | Peptide | Gamma-glutamyl Amino Acid | gamma-glutamyltyrosine | HMDB0011741 |
| grey | Cofactors and Vitamins | Tocopherol Metabolism | alpha-tocopherol | HMDB0001893 |
| grey | Amino Acid | Glutathione Metabolism | 2-aminobutyrate | HMDB0000452 |
| grey | Peptide | Gamma-glutamyl Amino Acid | gamma-glutamylglutamine | HMDB0011738 |
| grey | Amino Acid | Tyrosine Metabolism | 4-hydroxyphenylpyruvate | HMDB0000707 |
| grey | Lipid | Short Chain Fatty Acid | butyrate/isobutyrate (4:0) | HMDB0000039 |
| grey | Amino Acid | Creatine Metabolism | creatine | HMDB0000064 |
| grey | Amino Acid | Tyrosine Metabolism | 3-methoxytyrosine | HMDB0001434 |
| grey | Lipid | Primary Bile Acid Metabolism | glycochenodeoxycholate | HMDB0000637 |
| grey | Lipid | Primary Bile Acid Metabolism | taurochenodeoxycholate | HMDB0000951 |
| grey | Lipid | Primary Bile Acid Metabolism | taurocholate | HMDB0000036 |
| grey | Xenobiotics | Benzoate Metabolism | 2-hydroxyhippurate (salicylurate) | HMDB0000840 |
| grey | Lipid | Carnitine Metabolism | carnitine | HMDB0000062 |
| grey | Xenobiotics | Benzoate Metabolism | hippurate | HMDB0000714 |
| grey | Amino Acid | Tryptophan Metabolism | xanthurenate | HMDB0000881 |
| grey | Amino Acid | Leucine, Isoleucine and Valine Metabolism | 3-methyl-2-oxovalerate | HMDB0000491 |
| grey | Amino Acid | Methionine, Cysteine, SAM and Taurine Metabolism | methionine sulfoxide | HMDB0002005 |
| grey | Amino Acid | Histidine Metabolism | 3-methylhistidine | HMDB0000479 |
| grey | Amino Acid | Lysine Metabolism | 5-hydroxylysine | HMDB0000450 |
| grey | Lipid | Glycerolipid Metabolism | glycerol 3-phosphate | HMDB0000126 |
| grey | Lipid | Phospholipid Metabolism | glycerophosphorylcholine (GPC) | HMDB0000086 |
| grey | Amino Acid | Glutamate Metabolism | N-acetylglutamate | HMDB0001138 |
| grey | Carbohydrate | Pentose Metabolism | ribitol | HMDB0002917,HMDB0001851,HMDB0000568,HMDB0000508 |
| grey | Amino Acid | Tryptophan Metabolism | indolelactate | HMDB0000671 |
| grey | Amino Acid | Tryptophan Metabolism | 3-indoxyl sulfate | HMDB0000682 |
| grey | Amino Acid | Leucine, Isoleucine and Valine Metabolism | 4-methyl-2-oxopentanoate | HMDB0000695 |
| grey | Lipid | Sphingosines | sphingosine 1-phosphate | HMDB0000277 |
| grey | Lipid | Phosphatidylserine (PS) | 1-stearoyl-2-oleoyl-GPS (18:0/18:1) | HMDB0010163 |
| grey | Lipid | Lysophospholipid | 1-stearoyl-GPI (18:0) | HMDB0240261 |
| grey | Xenobiotics | Benzoate Metabolism | 4-acetylphenol sulfate | |
| grey | Lipid | Fatty Acid, Monohydroxy | 2-hydroxyoctanoate | HMDB0002264 |
| grey | Amino Acid | Phenylalanine Metabolism | phenyllactate (PLA) | HMDB0000779,HMDB0000563 |
| grey | Lipid | Androgenic Steroids | dehydroepiandrosterone sulfate (DHEA-S) | HMDB0001032 |
| grey | Amino Acid | Methionine, Cysteine, SAM and Taurine Metabolism | cysteine s-sulfate | HMDB0000731 |
| grey | Xenobiotics | Food Component/Plant | tartronate (hydroxymalonate) | HMDB0035227 |
| grey | Cofactors and Vitamins | Ascorbate and Aldarate Metabolism | oxalate (ethanedioate) | HMDB0002329 |
| grey | Xenobiotics | Chemical | iminodiacetate (IDA) | HMDB0011753 |

| | | | | |
|---|---|---|---|---|
| grey | Amino Acid | Leucine, Isoleucine and Valine Metabolism | 3-methyl-2-oxobutyrate | HMDB0000019 |
| grey | Amino Acid | Urea cycle; Arginine and Proline Metabolism | homoarginine | HMDB0000670 |
| grey | Lipid | Monoacylglycerol | 2-linoleoylglycerol (18:2) | HMDB0011538 |
| grey | Xenobiotics | Chemical | EDTA | HMDB0015109 |
| grey | Cofactors and Vitamins | Ascorbate and Aldarate Metabolism | threonate | HMDB0062620,HMDB0000943 |
| grey | Amino Acid | Tryptophan Metabolism | indoleacetate | HMDB0000197 |
| grey | Amino Acid | Leucine, Isoleucine and Valine Metabolism | isobutyrylcarnitine (C4) | HMDB0000736 |
| grey | Cofactors and Vitamins | Nicotinate and Nicotinamide Metabolism | trigonelline (N'-methylnicotinate) | HMDB0000875 |
| grey | Amino Acid | Tyrosine Metabolism | N-acetyltyrosine | HMDB0000866 |
| grey | Lipid | Fatty Acid Metabolism (also BCAA Metabolism) | propionylcarnitine (C3) | HMDB0000824 |
| grey | Amino Acid | Urea cycle; Arginine and Proline Metabolism | pro-hydroxy-pro | HMDB0006695 |
| grey | Lipid | Fatty Acid, Dicarboxylate | 3-carboxy-4-methyl-5-propyl-2-furanpropanoate (CMPF) | HMDB0061112 |
| grey | Amino Acid | Glutamate Metabolism | N-acetylglutamine | HMDB0006029 |
| grey | Amino Acid | Tryptophan Metabolism | N-acetyltryptophan | HMDB0013713 |
| grey | Amino Acid | Alanine and Aspartate Metabolism | N-acetylasparagine | HMDB0006028 |
| grey | Amino Acid | Urea cycle; Arginine and Proline Metabolism | N-acetylarginine | HMDB0004620 |
| grey | Lipid | Lysophospholipid | 1-stearoyl-GPC (18:0) | HMDB0010384 |
| grey | Lipid | Lysophospholipid | 1-oleoyl-GPC (18:1) | HMDB0002815 |
| grey | Lipid | Secondary Bile Acid Metabolism | hyocholate | HMDB0000760 |
| grey | Amino Acid | Histidine Metabolism | N-acetylhistidine | HMDB0032055 |
| grey | Peptide | Gamma-glutamyl Amino Acid | gamma-glutamylglycine | HMDB0011667 |
| grey | Xenobiotics | Food Component/Plant | stachydrine | HMDB0004827 |
| grey | Amino Acid | Leucine, Isoleucine and Valine Metabolism | alpha-hydroxyisovalerate | HMDB0000407 |
| grey | Peptide | Gamma-glutamyl Amino Acid | gamma-glutamylmethionine | HMDB0029155 |
| grey | Peptide | Gamma-glutamyl Amino Acid | gamma-glutamylthreonine | HMDB0029159 |
| grey | Xenobiotics | Benzoate Metabolism | p-cresol sulfate | HMDB0011635 |
| grey | Cofactors and Vitamins | Hemoglobin and Porphyrin Metabolism | heme | HMDB0003178 |
| grey | Amino Acid | Leucine, Isoleucine and Valine Metabolism | isovalerylcarnitine (C5) | HMDB0000688 |
| grey | Lipid | Lysophospholipid | 1-linoleoyl-GPC (18:2) | HMDB0010386 |
| grey | Nucleotide | Pyrimidine Metabolism, Cytidine containing | N4-acetylcytidine | HMDB0005923 |
| grey | Peptide | Acetylated Peptides | phenylacetylglutamine | HMDB0006344 |
| grey | Xenobiotics | Benzoate Metabolism | 4-hydroxyhippurate | HMDB0013678 |
| grey | Amino Acid | Glutathione Metabolism | cysteine-glutathione disulfide | HMDB0000656 |
| grey | Nucleotide | Pyrimidine Metabolism, Uracil containing | 5-methyluridine (ribothymidine) | HMDB0000884 |
| grey | Nucleotide | Pyrimidine Metabolism, Cytidine containing | 3-methylcytidine | HMDB0240577 |
| grey | Amino Acid | Tyrosine Metabolism | phenol sulfate | HMDB0060015 |
| grey | Lipid | Lysophospholipid | 1-palmitoleoyl-GPC (16:1)* | HMDB0010383 |
| grey | Amino Acid | Leucine, Isoleucine and Valine Metabolism | 2-hydroxy-3-methylvalerate | HMDB0000317 |
| grey | Lipid | Lysophospholipid | 2-palmitoyl-GPC (16:0)* | HMDB0061702 |
| grey | Lipid | Lysophospholipid | 1-oleoyl-GPE (18:1) | HMDB0011506 |
| grey | Lipid | Lysophospholipid | 1-linoleoyl-GPE (18:2)* | HMDB0011507 |
| grey | Amino Acid | Leucine, Isoleucine and Valine Metabolism | beta-hydroxyisovaleroylcarnitine | |
| grey | Xenobiotics | Drug - Topical Agents | hydroquinone sulfate | HMDB0240263 |
| grey | Xenobiotics | Benzoate Metabolism | catechol sulfate | HMDB0059724 |
| grey | Lipid | Sterol | cholesterol sulfate | HMDB00653 |
| grey | Amino Acid | Glutamate Metabolism | N-acetyl-aspartyl-glutamate (NAAG) | HMDB0001067 |
| grey | Lipid | Phospholipid Metabolism | glycerophosphoethanolamine | HMDB0000114 |
| grey | Lipid | Lysophospholipid | 1-palmitoyl-GPI (16:0) | HMDB0061695 |
| grey | Lipid | Secondary Bile Acid Metabolism | glycolithocholate sulfate* | HMDB0002639 |
| grey | Lipid | Secondary Bile Acid Metabolism | taurolithocholate 3-sulfate | HMDB0002580 |
| grey | Carbohydrate | Fructose, Mannose and Galactose Metabolism | mannitol/sorbitol | HMDB0000247,HMDB0000765 |
| grey | Xenobiotics | Benzoate Metabolism | 4-vinylphenol sulfate | HMDB0062775 |
| grey | Xenobiotics | Benzoate Metabolism | 4-ethylphenylsulfate | HMDB0062551 |
| grey | Xenobiotics | Food Component/Plant | thymol sulfate | HMDB0062720 |
| grey | Lipid | Lysophospholipid | 1-oleoyl-GPI (18:1) | HMDB0061693 |
| grey | Xenobiotics | Benzoate Metabolism | o-cresol sulfate | HMDB0011635 |
| grey | Xenobiotics | Food Component/Plant | 4-allylphenol sulfate | |
| grey | Lipid | Sphingolipid Synthesis | sphinganine-1-phosphate | HMDB0001383 |
| grey | Amino Acid | Urea cycle; Arginine and Proline Metabolism | N-methylproline | HMDB0094696 |
| grey | Lipid | Progestin Steroids | 5alpha-pregnan-3beta,20alpha-diol disulfate | HMDB0094650 |
| grey | Lipid | Secondary Bile Acid Metabolism | glycocholenate sulfate* | |
| grey | Lipid | Androgenic Steroids | androstenediol (3beta,17beta) disulfate (1) | HMDB0240313 |
| grey | Lipid | Pregnenolone Steroids | pregnenediol disulfate (C21H34O8S2)* | |
| grey | Lipid | Androgenic Steroids | androstenediol (3beta,17beta) disulfate (2) | HMDB0240313 |
| grey | Lipid | Pregnenolone Steroids | 21-hydroxypregnenolone disulfate | |
| grey | Lipid | Pregnenolone Steroids | pregnenediol sulfate (C21H34O5S)* | HMDB0000774 |
| grey | Cofactors and Vitamins | Tocopherol Metabolism | gamma-CEHC | HMDB0001931 |
| grey | Lipid | Androgenic Steroids | 16alpha-hydroxy DHEA 3-sulfate | HMDB0062544 |
| grey | Lipid | Pregnenolone Steroids | pregnenolone sulfate | HMDB0000774 |
| grey | Lipid | Androgenic Steroids | andro steroid monosulfate C19H28O6S (1)* | HMDB0002759 |
| grey | Amino Acid | Tryptophan Metabolism | indole-3-carboxylate | HMDB0003320 |
| grey | Xenobiotics | Food Component/Plant | 2,3-dihydroxyisovalerate | HMDB0012141 |
| grey | Lipid | Secondary Bile Acid Metabolism | isoursodeoxycholate | HMDB0000686 |
| grey | Amino Acid | Histidine Metabolism | formiminoglutamate | HMDB0000854 |
| grey | Amino Acid | Glutamate Metabolism | 4-hydroxyglutamate | HMDB0001344 |
| grey | Cofactors and Vitamins | Pantothenate and CoA Metabolism | pantoate | HMDB0240389 |
| grey | Amino Acid | Urea cycle; Arginine and Proline Metabolism | argininate* | HMDB0003148 |
| grey | Amino Acid | Urea cycle; Arginine and Proline Metabolism | 2-oxoarginine* | HMDB0004225 |
| grey | Lipid | Lysophospholipid | 1-lignoceroyl-GPC (24:0) | HMDB0010405 |
| grey | Amino Acid | Histidine Metabolism | 1-methyl-5-imidazoleacetate | HMDB04988 |
| grey | Lipid | Secondary Bile Acid Metabolism | glycoursodeoxycholate | HMDB0000708 |
| grey | Amino Acid | Methionine, Cysteine, SAM and Taurine Metabolism | S-methylcysteine sulfoxide | HMDB0029432 |
| grey | Lipid | Fatty Acid, Dicarboxylate | eicosanedioate (C20-DC) | |
| grey | Lipid | Fatty Acid, Dicarboxylate | docosadioate (C22-DC) | |
| grey | Peptide | Dipeptide | isoleucylglycine | HMDB0028907 |
| grey | Peptide | Dipeptide | valylleucine | HMDB0029131 |
| grey | Amino Acid | Glutamate Metabolism | beta-citrylglutamate | |
| grey | Lipid | Phospholipid Metabolism | trimethylamine N-oxide | HMDB0000925 |
| grey | Amino Acid | Lysine Metabolism | N6-methyllysine | HMDB0002038 |
| grey | Amino Acid | Histidine Metabolism | imidazole propionate | HMDB0002271 |
| grey | Peptide | Dipeptide | phenylalanylglycine | HMDB0028995 |
| grey | Peptide | Dipeptide | valylglutamine | HMDB0029125 |
| grey | Peptide | Dipeptide | valylglycine | HMDB0029127 |
| grey | Lipid | Lysophospholipid | 2-stearoyl-GPE (18:0)* | HMDB0011129 |
| grey | Lipid | Secondary Bile Acid Metabolism | glycohyocholate | HMDB0000138 |
| grey | Lipid | Fatty Acid, Monohydroxy | 2-hydroxydecanoate | HMDB0094656 |
| grey | Xenobiotics | Benzoate Metabolism | 4-methylcatechol sulfate | HMDB0240459 |
| grey | Xenobiotics | Benzoate Metabolism | 3-methyl catechol sulfate (1) | |
| grey | Xenobiotics | Benzoate Metabolism | guaiacol sulfate | HMDB0060013 |
| grey | Cofactors and Vitamins | Tocopherol Metabolism | gamma-CEHC glucuronide* | |
| grey | Xenobiotics | Food Component/Plant | 2-piperidinone | HMDB0011749 |
| grey | Xenobiotics | Food Component/Plant | 2-aminophenol sulfate | HMDB0061116 |
| grey | Amino Acid | Urea cycle; Arginine and Proline Metabolism | N-delta-acetylornithine | HMDB0003357 |
| grey | Amino Acid | Polyamine Metabolism | acisoga | HMDB0061384 |
| grey | Lipid | Fatty Acid, Amino | 2-aminoheptanoate | HMDB0094649 |
| grey | Amino Acid | Tyrosine Metabolism | 3-methoxytyramine sulfate | |
| grey | Lipid | Lysophospholipid | 1-linolenoyl-GPC (18:3)* | HMDB0010388 |
| grey | Amino Acid | Lysine Metabolism | fructosyllysine | HMDB0034879 |
| grey | Amino Acid | Leucine, Isoleucine and Valine Metabolism | 3-methylglutarylcarnitine (2) | HMDB0000552 |
| grey | Xenobiotics | Food Component/Plant | 2-keto-3-deoxy-gluconate | HMDB0001353 |
| grey | Amino Acid | Tyrosine Metabolism | N-formylphenylalanine | HMDB0240317 |
| grey | Xenobiotics | Chemical | 3-hydroxypyridine sulfate | |

| | | | | |
|---|---|---|---|---|
| grey | Peptide | Acetylated Peptides | phenylacetylcarnitine | |
| grey | Xenobiotics | Benzoate Metabolism | methyl-4-hydroxybenzoate sulfate | HMDB0041646 |
| grey | Xenobiotics | Benzoate Metabolism | propyl 4-hydroxybenzoate | HMDB0032574 |
| grey | Xenobiotics | Chemical | 6-hydroxyindole sulfate | HMDB0000682 |
| grey | Amino Acid | Tyrosine Metabolism | 4-methoxyphenol sulfate | |
| grey | Xenobiotics | Benzoate Metabolism | propyl 4-hydroxybenzoate sulfate | HMDB0135261 |
| grey | Xenobiotics | Food Component/Plant | umbelliferone sulfate | HMDB0240565 |
| grey | Lipid | Sphingomyelins | sphingomyelin (d18:1/20:2, d18:2/20:1, d16:1/22:2)* | |
| grey | Amino Acid | Tyrosine Metabolism | dopamine 3-O-sulfate | HMDB0006275 |
| grey | Xenobiotics | Benzoate Metabolism | 3-methoxycatechol sulfate (2) | |
| grey | Amino Acid | Tryptophan Metabolism | N-acetylkynurenine (2) | HMDB0240342 |
| grey | Lipid | Primary Bile Acid Metabolism | glycochenodeoxycholate 3-sulfate | HMDB0002409,HMDB0002496,HMDB0002497 |
| grey | Xenobiotics | Drug - Neurological | rocuronium | HMDB0014866 |
| grey | Lipid | Phosphatidylcholine (PC) | 1,2-dilinoleoyl-GPC (18:2/18:2) | HMDB0008138 |
| grey | Lipid | Phosphatidylcholine (PC) | 1-stearoyl-2-oleoyl-GPC (18:0/18:1) | HMDB0008038 |
| grey | Lipid | Phosphatidylcholine (PC) | 1-palmitoyl-2-docosahexaenoyl-GPC (16:0/22:6) | HMDB0007991 |
| grey | Lipid | Phosphatidylcholine (PC) | 1-stearoyl-2-docosahexaenoyl-GPC (18:0/22:6) | HMDB0008057 |
| grey | Lipid | Monoacylglycerol | 1-palmitoleoylglycerol (16:1)* | HMDB0011565 |
| grey | Lipid | Sphingomyelins | sphingomyelin (d18:2/24:1, d18:1/24:2)* | HMDB0240636,HMDB0240615 |
| grey | Lipid | Phosphatidylcholine (PC) | 1-stearoyl-2-linoleoyl-GPC (18:0/18:2)* | HMDB0008039 |
| grey | Lipid | Phosphatidylinositol (PI) | 1-stearoyl-2-linoleoyl-GPI (18:0/18:2) | HMDB0009809 |
| grey | Cofactors and Vitamins | Tocopherol Metabolism | gamma-tocopherol/beta-tocopherol | HMDB0006335,HMDB0001492 |
| grey | Lipid | Plasmalogen | 1-(1-enyl-palmitoyl)-2-oleoyl-GPE (P-16:0/18:1)* | HMDB0011342 |
| grey | Lipid | Plasmalogen | 1-(1-enyl-palmitoyl)-2-oleoyl-GPC (P-16:0/18:1)* | HMDB0007996 |
| grey | Lipid | Plasmalogen | 1-(1-enyl-palmitoyl)-2-linoleoyl-GPC (P-16:0/18:2)* | HMDB0011211 |
| grey | Lipid | Lactosylceramides (LCER) | lactosyl-N-palmitoyl-sphingosine (d18:1/16:0) | HMDB0006750 |
| grey | Amino Acid | Tryptophan Metabolism | 5-hydroxyindole sulfate | |
| grey | Lipid | Phosphatidylinositol (PI) | 1-palmitoyl-2-oleoyl-GPI (16:0/18:1)* | HMDB0009783 |
| grey | Lipid | Plasmalogen | 1-(1-enyl-palmitoyl)-2-linoleoyl-GPE (P-16:0/18:2)* | HMDB0011343 |
| grey | Lipid | Lysophospholipid | 1-linoleoyl-GPA (18:2)* | HMDB0007856 |
| grey | Lipid | Phosphatidylcholine (PC) | 1-oleoyl-2-docosahexaenoyl-GPC (18:1/22:6)* | HMDB0008123 |
| grey | Lipid | Phosphatidylcholine (PC) | 1-linoleoyl-2-arachidonoyl-GPC (18:2/20:4n6)* | HMDB0008147 |
| grey | Lipid | Phosphatidylcholine (PC) | 1-myristoyl-2-arachidonoyl-GPC (14:0/20:4)* | HMDB0007883 |
| grey | Lipid | Plasmalogen | 1-(1-enyl-palmitoyl)-2-palmitoleoyl-GPC (P-16:0/16:1)* | HMDB0011207 |
| grey | Lipid | Phosphatidylinositol (PI) | 1-stearoyl-2-oleoyl-GPI (18:0/18:1)* | HMDB0240667 |
| grey | Lipid | Lysophospholipid | 1-linoleoyl-GPG (18:2)* | HMDB0240600 |
| grey | Lipid | Fatty Acid Metabolism (Acyl Choline) | palmitoylcholine | HMDB0240592 |
| grey | Cofactors and Vitamins | Ascorbate and Aldarate Metabolism | ascorbic acid 2-sulfate | HMDB0060649 |
| grey | Lipid | Fatty Acid Metabolism (Acyl Choline) | arachidonoylcholine | HMDB0240583 |
| grey | Lipid | Phosphatidylcholine (PC) | 1-linoleoyl-2-linolenoyl-GPC (18:2/18:3)* | HMDB0008141 |
| grey | Lipid | Phosphatidylcholine (PC) | 1-palmitoleoyl-2-linolenoyl-GPC (16:1/18:3)* | HMDB0008008 |
| grey | Peptide | Gamma-glutamyl Amino Acid | gamma-glutamyl-alpha-lysine | |
| grey | Lipid | Lactosylceramides (LCER) | lactosyl-N-nervonoyl-sphingosine (d18:1/24:1)* | HMDB0004872 |
| grey | Lipid | Hexosylceramides (HCER) | glycosyl-N-(2-hydroxynervonoyl)-sphingosine (d18:1/24:1(2OH))* | |
| grey | Lipid | Fatty Acid Metabolism (Acyl Choline) | linoleoylcholine* | HMDB0013213 |
| grey | Lipid | Sphingomyelins | sphingomyelin (d18:2/24:2)* | HMDB0240644 |
| grey | Amino Acid | Polyamine Metabolism | (N(1) + N(8))-acetylspermidine | HMDB0002189,HMDB0001276 |
| grey | Lipid | Fatty Acid, Dicarboxylate | hydroxy-CMPF* | |
| grey | Carbohydrate | Pentose Metabolism | lyxonate | HMDB0060255 |
| grey | Lipid | Fatty Acid, Dicarboxylate | 3-carboxy-4-methyl-5-pentyl-2-furanpropionate (3-CMPFP)** | HMDB0061643 |
| grey | Lipid | Fatty Acid, Amino | N-acetyl-2-aminooctanoate* | HMDB0059745 |
| grey | Xenobiotics | Chemical | perfluorooctanoate (PFOA) | HMDB0059587 |
| grey | Xenobiotics | Food Component/Plant | 3-formylindole | HMDB29737 |
| grey | Peptide | Gamma-glutamyl Amino Acid | gamma-glutamylcitrulline* | |
| grey | Partially Characterized Molecules | Partially Characterized Molecules | glycine conjugate of C10H14O2 (1)* | |
| grey | Xenobiotics | Food Component/Plant | ethyl beta-glucopyranoside | HMDB0029968 |
| grey | Cofactors and Vitamins | Tocopherol Metabolism | delta-CEHC | |
| grey | Amino Acid | Lysine Metabolism | N6,N6-dimethyllysine | HMDB0013287 |
| grey | Amino Acid | Tryptophan Metabolism | indoleacetoylcarnitine* | |
| grey | Xenobiotics | Chemical | 2-naphthol sulfate | |
| grey | Amino Acid | Lysine Metabolism | N2-acetyl,N6,N6-dimethyllysine | |
| grey | Amino Acid | Lysine Metabolism | N2-acetyl,N6-methyllysine | |
| grey | Lipid | Secondary Bile Acid Metabolism | deoxycholic acid glucuronide | |
| grey | Xenobiotics | Food Component/Plant | methyl vanillate sulfate | |
| grey | Lipid | Secondary Bile Acid Metabolism | glycoursodeoxycholic acid sulfate (1) | |
| grey | Amino Acid | Leucine, Isoleucine and Valine Metabolism | 2-ketocaprylate | HMDB13211 |
| grey | Xenobiotics | Drug - Topical Agents | 2,6-dihydroxybenzoic acid | HMDB0013676 |
| grey | Lipid | Sphingomyelins | hydroxypalmitoyl sphingomyelin (d18:1/16:0(OH))** | |
| grey | Lipid | Secondary Bile Acid Metabolism | taurochenodeoxycholic acid 3-sulfate | HMDB0002486 |
| grey | Lipid | Pregnenolone Steroids | pregnenetriol sulfate* | |
| grey | Lipid | Pregnenolone Steroids | pregnenetriol disulfate* | |
| grey | Lipid | Fatty Acid Metabolism (Acyl Glycine) | picolinoylglycine | HMDB0059766 |
| grey | Peptide | Dipeptide | phenylalanylhydroxyproline* | HMDB0011176 |
| grey | Peptide | Dipeptide Derivative | leucylhydroxyproline* | HMDB0028930 |
| grey | Peptide | Dipeptide Derivative | isoleucylhydroxyproline* | HMDB0028908 |
| grey | Xenobiotics | Food Component/Plant | vanillic acid glycine | HMDB0060026 |
| grey | Amino Acid | Methionine, Cysteine, SAM and Taurine Metabolism | 2-hydroxy-4-(methylthio)butanoic acid | HMDB0037115 |
| grey | Amino Acid | Histidine Metabolism | 1-methyl-5-imidazolelactate | |
| grey | Amino Acid | Methionine, Cysteine, SAM and Taurine Metabolism | S-carboxyethylcysteine | |
| grey | Partially Characterized Molecules | Partially Characterized Molecules | carnitine of C10H14O2 (5)* | |
| grey | Amino Acid | Lysine Metabolism | N-acetyl-2-aminoadipate | |
| grey | Lipid | Fatty Acid Metabolism (Acyl Glycine) | cis-3,4-methyleneheptanoylglycine | |
| grey | Partially Characterized Molecules | Partially Characterized Molecules | bilirubin degradation product, C16H18N2O5 (2)** | |
| grey | Partially Characterized Molecules | Partially Characterized Molecules | bilirubin degradation product, C17H18N2O4 (1)** | |
| grey | Partially Characterized Molecules | Partially Characterized Molecules | bilirubin degradation product, C17H18N2O4 (2)** | |
| grey | Partially Characterized Molecules | Partially Characterized Molecules | bilirubin degradation product, C17H18N2O4 (3)** | |
| grey | Partially Characterized Molecules | Partially Characterized Molecules | bilirubin degradation product, C17H20N2O5 (2)** | |
| grey | Partially Characterized Molecules | Partially Characterized Molecules | bilirubin degradation product, C16H18N2O5 (3)** | |
| grey | Amino Acid | Tryptophan Metabolism | oxindolylalanine | |
| grey | Xenobiotics | Chemical | perfluorohexanesulfonate (PFHxS) | HMDB0094700 |
| grey | | | X-07765 | |
| grey | | | X-11299 | |
| grey | | | X-11308 | |
| grey | | | X-11315 | |
| grey | | | X-11372 | |
| grey | | | X-11444 | |
| grey | | | X-11470 | |
| grey | | | X-11880 | |
| grey | | | X-12007 | |
| grey | | | X-12112 | |
| grey | | | X-12193 | |
| grey | | | X-12216 | |
| grey | | | X-12410 | |
| grey | | | X-12456 | |
| grey | | | X-12462 | |
| grey | | | X-12729 | |
| grey | | | X-12812 | |
| grey | | | X-12844 | |
| grey | | | X-12906 | |
| grey | | | X-13431 | |
| grey | | | X-13684 | |
| grey | | | X-13729 | |

| | | | | |
|---|---|---|---|---|
| grey | | | X-13866 | |
| grey | | | X-15674 | |
| grey | | | X-16935 | |
| grey | | | X-16938 | |
| grey | | | X-17010 | |
| grey | | | X-17357 | |
| grey | | | X-18779 | |
| grey | | | X-18899 | |
| grey | | | X-19141 | |
| grey | | | X-21258 | |
| grey | | | X-21286 | |
| grey | | | X-21310 | |
| grey | | | X-21339 | |
| grey | | | X-21383 | |
| grey | | | X-21410 | |
| grey | | | X-21467 | |
| grey | | | X-21471 | |
| grey | | | X-21742 | |
| grey | | | X-21796 | |
| grey | | | X-21816 | |
| grey | | | X-22771 | |
| grey | | | X-23276 | |
| grey | | | X-23636 | |
| grey | | | X-23639 | |
| grey | | | X-23644 | |
| grey | | | X-23654 | |
| grey | | | X-23739 | |
| grey | | | X-24306 | |
| grey | | | X-24307 | |
| grey | | | X-24328 | |
| grey | | | X-24541 | |
| grey | | | X-24544 | |
| grey | | | X-24549 | |
| grey | | | X-24556 | |
| grey | | | X-24576 | |
| grey | | | X-24578 | |
| grey | | | X-24951 | |
| grey | | | X-24970 | |
| grey | | | X-25172 | |
| grey | | | X-25267 | |
| grey | | | X-25343 | |
| grey | | | X-25433 | |
| grey | | | X-25519 | |
| grey | | | X-25790 | |
| grey | | | X-26054 | |
| grey | | | X-26097 | |
| grey | | | X-26119 | |
| turquoise | Amino Acid | Tryptophan Metabolism | 5-hydroxyindoleacetate | HMDB0000763 |
| turquoise | Amino Acid | Tryptophan Metabolism | kynurenate | HMDB0000715 |
| turquoise | Amino Acid | Tyrosine Metabolism | homovanillate (HVA) | HMDB0000118 |
| turquoise | Amino Acid | Phenylalanine Metabolism | 4-hydroxyphenylacetate | HMDB0000020 |
| turquoise | Cofactors and Vitamins | Nicotinate and Nicotinamide Metabolism | quinolinate | HMDB0000232 |
| turquoise | Amino Acid | Lysine Metabolism | N6,N6,N6-trimethyllysine | HMDB0001325 |
| turquoise | Amino Acid | Polyamine Metabolism | N-acetylputrescine | HMDB0002064 |
| turquoise | Amino Acid | Methionine, Cysteine, SAM and Taurine Metabolism | N-formylmethionine | HMDB0001015 |
| turquoise | Amino Acid | Polyamine Metabolism | 5-methylthioadenosine (MTA) | HMDB0001173 |
| turquoise | Amino Acid | Phenylalanine Metabolism | 2-hydroxyphenylacetate | HMDB0000669 |
| turquoise | Amino Acid | Creatine Metabolism | creatinine | HMDB0000562 |
| turquoise | Cofactors and Vitamins | Vitamin A Metabolism | retinol (vitamin A) | HMDB0000305 |
| turquoise | Amino Acid | Urea cycle; Arginine and Proline Metabolism | urea | HMDB0000294 |
| turquoise | Nucleotide | Pyrimidine Metabolism, Uracil containing | pseudouridine | HMDB0000767 |
| turquoise | Nucleotide | Pyrimidine Metabolism, Cytidine containing | cytidine | HMDB0000089 |
| turquoise | Nucleotide | Pyrimidine Metabolism, Orotate containing | dihydroorotate | HMDB03349 |
| turquoise | Nucleotide | Purine Metabolism, (Hypo)Xanthine/Inosine containing | allantoin | HMDB0000462 |
| turquoise | Cofactors and Vitamins | Pantothenate and CoA Metabolism | pantothenate | HMDB0000210 |
| turquoise | Amino Acid | Methionine, Cysteine, SAM and Taurine Metabolism | N-acetylmethionine | HMDB0011745 |
| turquoise | Amino Acid | Leucine, Isoleucine and Valine Metabolism | N-acetylvaline | HMDB0011757 |
| turquoise | Amino Acid | Alanine and Aspartate Metabolism | N-acetylalanine | HMDB0000766 |
| turquoise | Amino Acid | Tyrosine Metabolism | vanillylmandelate (VMA) | HMDB0000291 |
| turquoise | Amino Acid | Polyamine Metabolism | 4-acetamidobutanoate | HMDB0003681 |
| turquoise | Nucleotide | Purine Metabolism, (Hypo)Xanthine/Inosine containing | urate | HMDB0000289 |
| turquoise | Carbohydrate | Aminosugar Metabolism | N-acetylneuraminate | HMDB0000230 |
| turquoise | Carbohydrate | Aminosugar Metabolism | N-acetylglucosaminylasparagine | HMDB0000489 |
| turquoise | Amino Acid | Glutathione Metabolism | cys-gly, oxidized | |
| turquoise | Nucleotide | Purine Metabolism, Adenine containing | N1-methyladenosine | HMDB0003331 |
| turquoise | Lipid | Phospholipid Metabolism | choline | HMDB0000097 |
| turquoise | Peptide | Gamma-glutamyl Amino Acid | gamma-glutamylleucine | HMDB0011171 |
| turquoise | Amino Acid | Histidine Metabolism | imidazole lactate | HMDB0002320 |
| turquoise | Amino Acid | Tryptophan Metabolism | kynurenine | HMDB0000684 |
| turquoise | Peptide | Gamma-glutamyl Amino Acid | gamma-glutamylphenylalanine | HMDB0000594 |
| turquoise | Xenobiotics | Food Component/Plant | erythritol | HMDB0002994 |
| turquoise | Amino Acid | Urea cycle; Arginine and Proline Metabolism | homocitrulline | HMDB0000679 |
| turquoise | Amino Acid | Histidine Metabolism | 1-methylhistidine | HMDB0000001 |
| turquoise | Cofactors and Vitamins | Vitamin B6 Metabolism | pyridoxate | HMDB0000017 |
| turquoise | Peptide | Gamma-glutamyl Amino Acid | gamma-glutamylvaline | HMDB0011172 |
| turquoise | Amino Acid | Histidine Metabolism | 1-methyl-4-imidazoleacetate | HMDB0002820 |
| turquoise | Amino Acid | Phenylalanine Metabolism | N-acetylphenylalanine | HMDB0000512 |
| turquoise | Amino Acid | Glycine, Serine and Threonine Metabolism | N-acetylthreonine | HMDB0062557 |
| turquoise | Amino Acid | Leucine, Isoleucine and Valine Metabolism | N-acetylisoleucine | HMDB0061684 |
| turquoise | Carbohydrate | Aminosugar Metabolism | erythronate* | HMDB0000613 |
| turquoise | Nucleotide | Purine Metabolism, (Hypo)Xanthine/Inosine containing | N1-methylinosine | HMDB0002721 |
| turquoise | Nucleotide | Purine Metabolism, Guanine containing | N2,N2-dimethylguanosine | HMDB0004824 |
| turquoise | Nucleotide | Purine Metabolism, Adenine containing | N6-carbamoylthreonyladenosine | HMDB0041623 |
| turquoise | Nucleotide | Pyrimidine Metabolism, Orotate containing | orotidine | HMDB0000788 |
| turquoise | Nucleotide | Pyrimidine Metabolism, Uracil containing | 5,6-dihydrouridine | HMDB0000497 |
| turquoise | Nucleotide | Pyrimidine Metabolism, Uracil containing | 3-(3-amino-3-carboxypropyl)uridine* | |
| turquoise | Nucleotide | Purine Metabolism, Guanine containing | 7-methylguanine | HMDB0000897 |
| turquoise | Cofactors and Vitamins | Nicotinate and Nicotinamide Metabolism | N1-methyl-2-pyridone-5-carboxamide | HMDB0004193 |
| turquoise | Peptide | Gamma-glutamyl Amino Acid | gamma-glutamylisoleucine* | HMDB0011170 |
| turquoise | Amino Acid | Leucine, Isoleucine and Valine Metabolism | 2-methylbutyrylcarnitine (C5) | HMDB0000378 |
| turquoise | Partially Characterized Molecules | Partially Characterized Molecules | glutamine_degradant* | |
| turquoise | Cofactors and Vitamins | Ascorbate and Aldarate Metabolism | gulonate* | HMDB0003290 |
| turquoise | Amino Acid | Lysine Metabolism | glutarylcarnitine (C5-DC) | HMDB0013130 |
| turquoise | Amino Acid | Leucine, Isoleucine and Valine Metabolism | tiglylcarnitine (C5:1-DC) | HMDB0002366 |
| turquoise | Nucleotide | Purine Metabolism, Adenine containing | N6-succinyladenosine | HMDB0000912 |
| turquoise | Amino Acid | Histidine Metabolism | 1-ribosyl-imidazoleacetate* | HMDB0002331 |
| turquoise | Lipid | Fatty Acid, Dihydroxy | 3,4-dihydroxybutyrate | HMDB0000337 |
| turquoise | Amino Acid | Lysine Metabolism | N6-acetyllysine | HMDB0000206 |
| turquoise | Amino Acid | Glycine, Serine and Threonine Metabolism | N-acetylserine | HMDB0002931 |
| turquoise | Energy | TCA Cycle | succinylcarnitine (C4-DC) | HMDB0061717 |
| turquoise | Nucleotide | Pyrimidine Metabolism, Uracil containing | N-acetyl-beta-alanine | HMDB0061880 |
| turquoise | Lipid | Fatty Acid, Dihydroxy | 2R,3R-dihydroxybutyrate | HMDB0000498 |

| | | | | |
|---|---|---|---|---|
| turquoise | Amino Acid | Glutamate Metabolism | alpha-ketoglutaramate* | HMDB0001552 |
| turquoise | Amino Acid | Glutathione Metabolism | cysteinylglycine disulfide* | HMDB0000709 |
| turquoise | Cofactors and Vitamins | Ascorbate and Aldarate Metabolism | 2-O-methylascorbic acid | HMDB0240294 |
| turquoise | Xenobiotics | Food Component/Plant | mannonate* | |
| turquoise | Amino Acid | Methionine, Cysteine, SAM and Taurine Metabolism | lanthionine | |
| turquoise | Amino Acid | Histidine Metabolism | N-acetylcarnosine | HMDB0012881 |
| turquoise | Amino Acid | Urea cycle; Arginine and Proline Metabolism | N2,N5-diacetylornithine | HMDB0240345 |
| turquoise | Amino Acid | Methionine, Cysteine, SAM and Taurine Metabolism | methionine sulfone | HMDB0062174 |
| turquoise | Xenobiotics | Chemical | O-sulfo-tyrosine | HMDB0155722 |
| turquoise | Amino Acid | Methionine, Cysteine, SAM and Taurine Metabolism | N-acetyltaurine | HMDB0240253 |
| turquoise | Amino Acid | Tyrosine Metabolism | tyramine O-sulfate | HMDB0006409 |
| turquoise | Carbohydrate | Pentose Metabolism | arabonate/xylonate | HMDB0000539 |
| turquoise | Amino Acid | Tyrosine Metabolism | vanillactate | HMDB0000913 |
| turquoise | Amino Acid | Tryptophan Metabolism | C-glycosyltryptophan | HMDB0240296 |
| turquoise | Carbohydrate | Pentose Metabolism | arabitol/xylitol | HMDB0001851,HMDB0000568,HMDB0002917 |
| turquoise | Carbohydrate | Aminosugar Metabolism | N-acetylglucosamine/N-acetylgalactosamine | HMDB0000212,HMDB0000215 |
| turquoise | Peptide | Acetylated Peptides | 4-hydroxyphenylacetylglutamine | |
| turquoise | Amino Acid | Alanine and Aspartate Metabolism | hydroxyasparagine** | HMDB32332 |
| turquoise | Amino Acid | Urea cycle; Arginine and Proline Metabolism | N,N,N-trimethyl-alanylproline betaine (TMAP) | HMDB0240365 |
| turquoise | Amino Acid | Leucine, Isoleucine and Valine Metabolism | N-carbamoylvaline | |
| turquoise | Amino Acid | Urea cycle; Arginine and Proline Metabolism | 3-amino-2-piperidone | HMDB0000323 |
| turquoise | Cofactors and Vitamins | Ascorbate and Aldarate Metabolism | ascorbic acid 3-sulfate* | |
| turquoise | Amino Acid | Polyamine Metabolism | N-acetyl-isoputreanine | |
| turquoise | Amino Acid | Methionine, Cysteine, SAM and Taurine Metabolism | 2,3-dihydroxy-5-methylthio-4-pentenoate (DMTPA)* | HMDB0240388 |
| turquoise | Amino Acid | Lysine Metabolism | hydroxy-N6,N6,N6-trimethyllysine* | |
| turquoise | Peptide | Modified Peptides | N,N-dimethyl-pro-pro | |
| turquoise | | | X-11787 | |
| turquoise | | | X-11979 | |
| turquoise | | | X-12015 | |
| turquoise | | | X-12026 | |
| turquoise | | | X-12100 | |
| turquoise | | | X-12104 | |
| turquoise | | | X-12411 | |
| turquoise | | | X-12713 | |
| turquoise | | | X-12822 | |
| turquoise | | | X-13553 | |
| turquoise | | | X-15461 | |
| turquoise | | | X-15503 | |
| turquoise | | | X-17676 | |
| turquoise | | | X-18887 | |
| turquoise | | | X-22162 | |
| turquoise | | | X-24337 | |
| turquoise | | | X-24588 | |
| turquoise | | | X-25371 | |
| turquoise | | | X-25422 | |
| turquoise | | | X-25810 | |
| yellow | Lipid | Long Chain Polyunsaturated Fatty Acid (n3 and n6) | linoleate (18:2n6) | HMDB0006270,HMDB0000673 |
| yellow | Lipid | Medium Chain Fatty Acid | laurate (12:0) | HMDB0000638 |
| yellow | Lipid | Long Chain Polyunsaturated Fatty Acid (n3 and n6) | arachidonate (20:4n6) | HMDB0001043 |
| yellow | Lipid | Long Chain Saturated Fatty Acid | palmitate (16:0) | HMDB0000220 |
| yellow | Lipid | Long Chain Saturated Fatty Acid | stearate (18:0) | HMDB0000827 |
| yellow | Lipid | Long Chain Monounsaturated Fatty Acid | palmitoleate (16:1n7) | HMDB0003229 |
| yellow | Lipid | Long Chain Saturated Fatty Acid | myristate (14:0) | HMDB0000806 |
| yellow | Lipid | Medium Chain Fatty Acid | caprate (10:0) | HMDB0000511 |
| yellow | Lipid | Long Chain Saturated Fatty Acid | margarate (17:0) | HMDB0002259 |
| yellow | Lipid | Long Chain Saturated Fatty Acid | nonadecanoate (19:0) | HMDB0000772 |
| yellow | Lipid | Long Chain Saturated Fatty Acid | arachidate (20:0) | HMDB0002212 |
| yellow | Lipid | Medium Chain Fatty Acid | caproate (6:0) | HMDB0000535 |
| yellow | Lipid | Long Chain Saturated Fatty Acid | pentadecanoate (15:0) | HMDB0000826 |
| yellow | Lipid | Long Chain Monounsaturated Fatty Acid | erucate (22:1n9) | HMDB0002068 |
| yellow | Lipid | Long Chain Polyunsaturated Fatty Acid (n3 and n6) | dihomo-linoleate (20:2n6) | HMDB0005060 |
| yellow | Lipid | Fatty Acid, Monohydroxy | 2-hydroxystearate | HMDB0062549 |
| yellow | Lipid | Glycerolipid Metabolism | glycerol | HMDB0000131 |
| yellow | Lipid | Long Chain Polyunsaturated Fatty Acid (n3 and n6) | eicosapentaenoate (EPA; 20:5n3) | HMDB0001999 |
| yellow | Lipid | Long Chain Polyunsaturated Fatty Acid (n3 and n6) | docosahexaenoate (DHA; 22:6n3) | HMDB0002183 |
| yellow | Lipid | Fatty Acid, Monohydroxy | 3-hydroxymyristate | HMDB0061656 |
| yellow | Lipid | Monoacylglycerol | 1-oleoylglycerol (18:1) | HMDB0011567 |
| yellow | Lipid | Fatty Acid, Monohydroxy | 3-hydroxydecanoate | HMDB0002203 |
| yellow | Lipid | Monoacylglycerol | 1-linoleoylglycerol (18:2) | HMDB0011568 |
| yellow | Lipid | Fatty Acid, Monohydroxy | 3-hydroxylaurate | HMDB0000387 |
| yellow | Amino Acid | Leucine, Isoleucine and Valine Metabolism | 3-hydroxy-2-ethylpropionate | HMDB0000396 |
| yellow | Lipid | Long Chain Polyunsaturated Fatty Acid (n3 and n6) | docosapentaenoate (n3 DPA; 22:5n3) | HMDB0006528,HMDB0001976 |
| yellow | Lipid | Long Chain Polyunsaturated Fatty Acid (n3 and n6) | docosadienoate (22:2n6) | HMDB0061714 |
| yellow | Lipid | Long Chain Polyunsaturated Fatty Acid (n3 and n6) | adrenate (22:4n6) | HMDB0002226 |
| yellow | Lipid | Long Chain Monounsaturated Fatty Acid | myristoleate (14:1n5) | HMDB0002000 |
| yellow | Lipid | Long Chain Polyunsaturated Fatty Acid (n3 and n6) | stearidonate (18:4n3) | HMDB0006547 |
| yellow | Lipid | Medium Chain Fatty Acid | 5-dodecenoate (12:1n7) | HMDB0000529 |
| yellow | Lipid | Long Chain Monounsaturated Fatty Acid | 10-nonadecenoate (19:1n9) | HMDB0013622 |
| yellow | Lipid | Long Chain Monounsaturated Fatty Acid | 10-heptadecenoate (17:1n7) | HMDB0060038 |
| yellow | Lipid | Long Chain Monounsaturated Fatty Acid | eicosenoate (20:1) | HMDB0002231,HMDB0062436 |
| yellow | Lipid | Long Chain Polyunsaturated Fatty Acid (n3 and n6) | linolenate [alpha or gamma; (18:3n3 or 6)] | HMDB0003073,HMDB0001388 |
| yellow | Lipid | Monoacylglycerol | 1-arachidonylglycerol (20:4) | HMDB11578 |
| yellow | Lipid | Fatty Acid, Monohydroxy | 2-hydroxypalmitate | HMDB0031057 |
| yellow | Lipid | Long Chain Polyunsaturated Fatty Acid (n3 and n6) | docosapentaenoate (n6 DPA; 22:5n6) | HMDB0001976 |
| yellow | Lipid | Sterol | 7alpha-hydroxy-3-oxo-4-cholestenoate (7-Hoca) | HMDB0012458 |
| yellow | Lipid | Lysophospholipid | 1-arachidonoyl-GPI (20:4)* | HMDB0061690 |
| yellow | Lipid | Long Chain Polyunsaturated Fatty Acid (n3 and n6) | dihomo-linolenate (20:3n3 or n6) | HMDB0002925 |
| yellow | Lipid | Lysophospholipid | 1-linoleoyl-GPI (18:2)* | HMDB0240597 |
| yellow | Lipid | Fatty Acid, Branched | (16 or 17)-methylstearate (a19:0 or i19:0) | HMDB0037397 |
| yellow | Lipid | Medium Chain Fatty Acid | cis-4-decenoate (10:1n6)* | HMDB0004980 |
| yellow | Lipid | Fatty Acid Metabolism (Acyl Glycine) | N-palmitoylglycine | HMDB0013034 |
| yellow | Lipid | Lysoplasmalogen | 1-(1-enyl-oleoyl)-GPE (P-18:1)* | |
| yellow | Lipid | Fatty Acid, Monohydroxy | 9-hydroxystearate | HMDB0061661 |
| yellow | Lipid | Sterol | 3beta-hydroxy-5-cholestenoate | HMDB0012453 |
| yellow | Lipid | Endocannabinoid | linoleoyl ethanolamide | HMDB0012252 |
| yellow | Lipid | Long Chain Monounsaturated Fatty Acid | oleate/vaccenate (18:1) | HMDB0003231,HMDB0000573,HMDB0240219,HMDB0000207 |
| yellow | Lipid | Long Chain Polyunsaturated Fatty Acid (n3 and n6) | hexadecadienoate (16:2n6) | HMDB0000477 |
| yellow | Lipid | Fatty Acid, Monohydroxy | 2-hydroxybehenate | HMDB0061660 |
| yellow | Lipid | Long Chain Polyunsaturated Fatty Acid (n3 and n6) | tetradecadienoate (14:2)* | HMDB0000560 |
| yellow | Lipid | Fatty Acid, Dicarboxylate | dodecadienoate (12:2)* | |
| yellow | Lipid | Medium Chain Fatty Acid | (2 or 3)-decenoate (10:1n7 or n8) | |
| yellow | Partially Characterized Molecules | Partially Characterized Molecules | branched-chain, straight-chain, or cyclopropyl 10:1 fatty acid (1)* | |
| yellow | Partially Characterized Molecules | Partially Characterized Molecules | branched-chain, straight-chain, or cyclopropyl 12:1 fatty acid* | |
| yellow | | | X-21353 | |
| yellow | | | X-23782 | |
